# Distinct gut and vaginal microbiota profile in women with Recurrent Implantation Failure and Unexplained Infertility

**DOI:** 10.1101/2021.02.09.21251410

**Authors:** Nayna Patel, Nidhi Patel, Sejal Pal, Neelam Nathani, Ramesh Pandit, Molina Patel, Niket Patel, Chaitanya Joshi, Bhavin Parekh

## Abstract

**Background:** Implantation failure limits the success rate of natural and *in vitro* fertilization (IVF)-assisted conceptions. Evidence suggests dysbiosis in the female reproductive tract impacts implantation failure. However, whether gut dysbiosis influences implantation failure and whether it accompanies reproductive tract dysbiosis remains unexplored.

**Method:** We recruited 11fertile women as the controls, and a cohort of 20 women diagnosed with implantation-failure associated infertility, which included 10 women diagnosed with recurrent implantation failure (RIF), and 10 women diagnosed with unexplained infertility (UE). Using next-generation amplicon sequencing, we compared the diversity, structure, and composition of fecal and vaginal bacteria of the controls with that of the infertile cohort. While we sequenced fecal samples of all the participants (n=31), we could only sequence 8 vaginal samples in each group (n=24).

**Results:** Compared with the controls, α-diversity of the gut bacteria, analysed by Chao 1 and Shannon indices, among the infertile groups declined (*p*□<□0.05). β-diversity between the controls and infertile cohort, measured by both Bray-Curtis and Jaccard distances, differed significantly (*p*□<□0.05). Taxa analysis of the gut bacteria revealed enrichment of Gram-positive bacteria, mainly of the phylum *Firmicutes*, in the RIF group. In contrast, Gram-negative bacteria were relatively more abundant in the UE group. Additionally, mucus-producing bacteria genera such as *Prevotella* and *Sutterella* declined in the infertile cohort (*p*□<□0.05). Intriguingly, significant enrichment (*p*□<□0.05) of the genus *Hungatella*, associated with trimethylamine N-oxide (TMAO) production, occurred in the infertile cohort. Vaginal microbiota was dominated by *L. iners* across the groups, with the UE group showing the highest levels. Of the three groups, the RIF group had the least diverse vaginal microbiota. Taxa analysis showed higher levels of anaerobic bacteria such as *Leptotrichia, Snethia*, and *Prevotella* in the controls.

**Conclusion:** We posit that in the setting of the compromised gut mucosal barrier, the phyla *Firmicutes* generates TNF-α*-*driven systemic inflammation, leading to RIF, whereas an overload of Gram-negative bacteria induces IL-6-driven systemic inflammation, leading to UE. Additionally, *Hungatella-*induced elevation of TMAO levels causes platelet hypercoagulability, synergistically contributing to implantation failure. Finally, vaginal dysbiosis does not appear to co-occur with gut dysbiosis.

## 1. Introduction

Infertility, defined as a failure to conceive after 1 year of appropriately timed unprotected intercourse, is a distressing and costly reproductive disorder that affects up to 15% of couples globally (Kamel 2010). Some known causes of infertility include pelvic diseases, peritoneal factors, cervical factors, ovulatory disorders, reproductive aging of women, and male factors (Lindsay and Vitrikas 2015).

Despite this knowledge, some couples are diagnosed as having unexplained infertility (UE) because the underlying mechanism(s) remain undefined even after assessment of ovulatory function, tubal patency, and sperm parameters (Bellver, Soares et al. 2008). Frustratingly, many of the infertile couples undergo multiple unsuccessful assisted reproduction technology (ART) cycles (i.e., IVF and/or intracytoplasmic sperm injection (ICSI)) and are thus diagnosed as having repeated implantation failure (RIF) (Bellver, Soares et al. 2008, Simon and Laufer 2012, Polanski, Baumgarten et al. 2014). Yet another subgroup of infertile couples, diagnosed as recurrent pregnancy loss (RPL), exists that conceive several times (≥3), but miscarriage occurs each time before gestational week 28, although controversies exist on its definition (Christiansen, Nielsen et al. 2006, El Hachem, Crepaux et al. 2017). It has been argued that RIF and RPL represent the same condition spectrum (Christiansen, Nielsen et al. 2006).

Pathologies of all these enigmatic conditions converge on mechanisms by which the embryo fails to implant in the uterus—also called implantation failure (Graham, Seif et al. 1990, Christiansen, Nielsen et al. 2006, Bellver, Soares et al. 2008, Simon and Laufer 2012). It is the main limiting factor for natural and *in vitro* fertilization (IVF)-assisted pregnancies (Bellver, Soares et al. 2008, Hernández-Vargas, Muñoz et al. 2020). Failed implantation involves a triumvirate of a poor quality embryo and an unreceptive endometrium and ill-timed embryo-endometrium interaction, of which unreceptive endometrium appears to be the most critical factor. (Simon and Laufer 2012). In fact, unreceptive endometrium has been implicated in two-thirds of all the failures (Simon and Laufer 2012). While multiple factors that disrupt endometrium receptivity, including steroidal hormonal imbalance, thrombotic abnormalities, hyperhomocysteinemia, and immune dysfunctions, have been identified, much remains recondite (Bellver, Soares et al. 2008, Simon and Laufer 2012, Cho, Kim et al. 2019).

The reproductive tract bacteria influence implantation failure of endometrial origin (Al-Nasiry, Ambrosino et al. 2020). For example, women with non-*Lactobacillus*-dominated microbiota in a receptive endometrium had decreased implantation rates than women with a *Lactobacillus*-dominated (>90%) microbiota (Moreno, Codoñer et al. 2016). Chlamydia species in the endocervix of women undergoing IVF-ET were correlated with implantation failure (Witkin, Sultan et al. 1994).

Increasingly researchers are exploring how the local microbiota influences physiology at distal sites, especially how the gut bacteria, the densest and most diverse bacterial communities of the body, impact distal organs, including the brain and lungs, leading to the notions of the gut-brain axis and gut–lung microbiota axis (Ravel and Brotman 2016, Budden, Gellatly et al. 2017). However, hitherto, gut bacteria’s role in implantation failure of endometrial origin remains unexplored, let alone the gut-reproductive tract microbiota axis, even though a compelling rationale exists. Indeed, the gut bacteria impact the immune system, hormonal homeostasis, and the coagulation system—all of which mediate implantation success (Alexander, Targan et al. 2014, Baker, Al-Nakkash et al. 2017, Vinchi 2019). The comorbidity of gut disorders (e.g., celiac disease) with infertility disorders, including recurrent pregnancy loss, allude to a role of ‘gut-reproductive tract axis’ in implantation failure (Tersigni, D’Ippolito et al. 2018).

Hence we investigated whether gut dysbiosis occurs in women with implantation failure, and if so, whether it accompanies vaginal dysbiosis, which usually involves a decline in either the levels of *Lactobacillus* or protective species of *Lactobacillus* and a simultaneous rise in the diversity and density of other bacteria (Madhivanan, Alleyn et al. 2015, Amabebe and Anumba 2018, Wang, Fan et al. 2020). To this end, by using 16S rRNA gene sequencing, we compared the diversity, structure and taxonomic composition of the fecal and vaginal microbiota of fertile women with that of infertile women with a history of RIF and UE.

## 2. Materials and Methods

### 2.1 Study participants

For this retrospective, single-center cohort study, fertile and infertile women, referred to Akanksha Hospital and Research Institute between September 2018 and February 2019, were recruited and divided into three groups: the control, RIF, and UE groups. The RIF group’s inclusion criteria were women who could not conceive after ≥2 fresh IVF-embryo transfer cycles/ ICSI or had ≥3 consecutive miscarriages. UE was diagnosed if a cause remains undefined after our routine fertility tests with the following criteria: infertility of more than 1 year, normal male partner, normal menstrual rhythm with regular ovulation, and normal hormonal tests (i.e., thyroid, prolactin, AMH) Exclusion criteria included diabetes, polycystic ovary syndrome and endometriosis, diarrhea, ongoing pregnancy, addiction (e.g., drugs, alcohol, tobacco etc.) and the use of antibiotics within at least two weeks before sample collection. We performed all the sampling and experiments with the approval of the local Ethics Committee of Sat Kaival Hospital Pvt. Ltd (EC2013/053). Participants gave their oral and written informed consent for fecal and vaginal sample collections and subsequent microbiological analysis. We recorded and compared participants’ characteristics **(Table 1)**.

**Table 1.** Study characteristics of the control, RIF, and the UE groups. Data are expressed as the mean ± SD or n/N (%).RIF, recurrent implantation failure; UE, unexplained infertility; BMI, body–mass index; Veg, Vegetarian; NA, Not applicable. Differences between the 3 groups were assessed by 1-factor ANOVA and Tukey’s multiple comparison test was used for post-hoc comparisons, if P<0.05. In the RIF group, two participants belonged to the RPL category. a. statistically significant difference between CON and RIF.

| Characteristics | Control<br>(n=11) |  | RIF<br>(n=10) |  | UE<br>(n=10) |  |
| --- | --- | --- | --- | --- | --- | --- |
| Age<br>(Years) | 27.9±3.8 |  | 34.5±4.8<sup>a</sup> |  | 30.8±3.42 |  |
| BMI<br>(kg/m<sup>2</sup>) | 21.62±4.05 |  | 25.9±3.31<sup>a</sup> |  | 23.92±3 |  |
| Nulligravida | NA |  | 2 (20%) |  | NA |  |
| Nullipara |  |  | 8 (80%) |  | 10 (100%) |  |
| Dietary Information | Veg | Non-Veg | Veg | Non-Veg | Veg | Non-Veg |
|  | 5<br>(45%) | 6<br>(54%) | 4<br>(40%) | 6<br>(60%) | 9<br>(90%) | 1<br>(10%) |

### 2.2 Sample Collection

The fecal and vaginal samples were freshly and simultaneously collected. Participants collected the fecal samples in a sterile plastic container with a tight closing lid. To collect the vaginal samples, using a sterile swab stick, clinicians thoroughly wiped the posterior fornix of the vagina of the participants. These swabs were stored in sterile vials. Both types of samples were packaged and first placed in a frozen storage at −20°C in the hospital and later, within 24 hours, transported on ice to be stored at −80°C at Gujarat Biotechnology Research Centre (GBRC) for further analysis.

### 2.3 DNA extraction

DNA extraction was performed from approximately 200 mg of fecal samples and ∼1ml of thoroughly vortexed swab sample using QIAamp DNA Stool Mini Kit according to the manufacturer’s instructions. Total DNA was eluted in 30 μL of AE buffer. DNA concentration was quantified fluorometrically with a Qubit 2.0 dsDNA HS Assay kit. DNA was stored at-20^0^C for further procedures.

### 2.4 Library preparation and 16s rRNA Sequencing

The V2-V3 hypervariable regions of the 16S rRNA gene were amplified using fusion primers, 101□F5′ACTGGCGGACGGGTGAGTAA 3′ and 518□R 5′CGTATTACCGCGGCTGCTGG 3′. Amplicon libraries were purified using the Agencourt AMPure XP (Beckman Coulter). For quality control, we used Bioanalyzer with a DNA-HS assay kit. All the libraries were quantified using Qubit fluorimeter v4.0 and were pooled into equimolar concentrations. Clonal amplification (Emulsion PCR) sequencing was performed on the Ion S5 system using the Ion 520/530 kit OT2 (ThermoFisher Scientific) with Ion One Touch 2 system, which uses a 530 chip on Ion S5 plus sequencer according to the manufacturer’s instructions.

### 2.5 Bioinformatics and Statistical analysis

#### Diversity

Microbial community richness and diversity was evaluated by α-diversity that included Chao1 and Shannon indices. The Kruskal–Wallis test used to determine statistical differences between the three groups. The Mann–Whitney U test was used to determine the influence of diet on α-diversity. β-diversity, differences in microbial community structures between the three groups, was analysed using Principle Coordinate Analysis (PCoA) based on Bray-Curtis and Jaccard distances, and the statistical difference between the groups was calculated using non-parametric multivariate analysis of variance (PERMANOVA).

QIIME2 software was used to calculate alpha and beta diversity indices. The demultiplexed sequences were uploaded to QIIME2 environment, and denoising was carried out using DADA2. Amplicon sequence variants (ASVs) were predicted at a minimum sampling depth of 25000 for Gut datasets, and 9000 for the vaginal datasets. The predicted ASVs were taxonomically classified using the pre-trained classifier of the full 16S rRNA gene sequence of the SILVA database.

#### Taxonomic structure

We analysed the gut microbiota at the community level to determine differences in the microbial composition between groups and to identify taxa with significantly different abundance (relative abundance > 0.001 and P < 0.05). Additionally, linear discriminant analysis (LDA) Effect Size (LEfSe) method was employed to identify species with significant differences in abundance between the three groups (|LDA| > 3 and P < 0.05). Kruskal–Wallis and Mann–Whitney U tests were performed for computing statistical differences between the groups at taxonomic levels in the study viz., three major groups and the infertile cohort versus controls using STAMP v2.1 software (Parks et al., 2014).

Continuous data are presented as mean ± standard deviation (SD) or frequencies (number and percentages), calculated using GraphPad Prism statistical software 6.0. To determine statistical differences in subjects’ characteristics between groups, we performed one-way ANOVA followed by post-hoc Tukey testing.

## 3. Results

### 3.1 Clinical Characteristics among the Control, RIF, and UE Groups

We included 31 study subjects, comprising 11 fertile women, enrolled as the control group, 10 women with a history of RIF, and 10 women with a history of UE **(Table 1)**. Comparisons of the mean age between the three groups showed that the controls were statistically significantly younger than the RIF group (F=6.8, p <0.01), while the UE group was older than the controls and younger than the RIF group, although both differences were statistically insignificant **(Table 1)**. The average BMI was not significantly different between the controls and the UE group and between the RIF and UE groups but was significant between the controls and the RIF group (F=3.7, p<0.05) **(Table 1)**. Among women with RIF, eight were nullipara (80%), and two were nulligravida (20%), while all women with UE (100%) were nullipara. There were more vegetarian participants in the UE group (9 of 10; 90%) than in the RIF group (4 of 10; 40%) and the controls (4 of11; 36%) **(Table 1)**.

### 3.2 Metagenomics Findings of Gut and Vaginal bacteria

In total 31 fecal samples and 24 vaginal swab samples, sequencing of the V2-V3 region of the 16S rRNA gene created 235810 sequences, with an average of 1,01, 767 sequences, and 178260 high-quality sequences, with an average of 3,76,012 sequences, per sample for fecal and vaginal samples respectively. Based on the results of the operational taxonomic units (OTUs) analysis, rarefaction curves show that the sequencing depth was adequate to analyse the gut and vaginal bacteria in the three groups **(Figures S1 (a), (b) and Figures S2 (a), (b))**.

### 3.3 Richness and diversity of Gut bacteria

We performed bacterial diversity analyses by comparing the richness using Chao 1 and evenness using the Shannon index for the fecal microbial communities between the three groups. We found that richness differed significantly between the three groups (Chao 1 index (Kruskal–Wallis test, *p*= 0.049)) **(Figure 1(a))**. Precisely, the controls had a significantly higher richness than the RIF group (pChao 1 = 0.04) and UE group (pChao 1 = 0.03). Richness was similar between RIF and UE groups (pChao 1 = 0.75). We discovered that evenness differed significantly between the three groups: Shannon index (Kruskal–Wallis test, *p* = 0.003) **(Figure 1(b))**. Specifically, the controls had more evenness than the RIF (pShannon = 0.006) and the UE groups (pShannon = 0.002). In contrast, evenness was similar between the RIF and the UE groups (pShannon = 0.65). Interestingly, the diet did not affect alpha diversity (pShannon = 0.165) **(Figure 3S)**.

**Figure 1.**
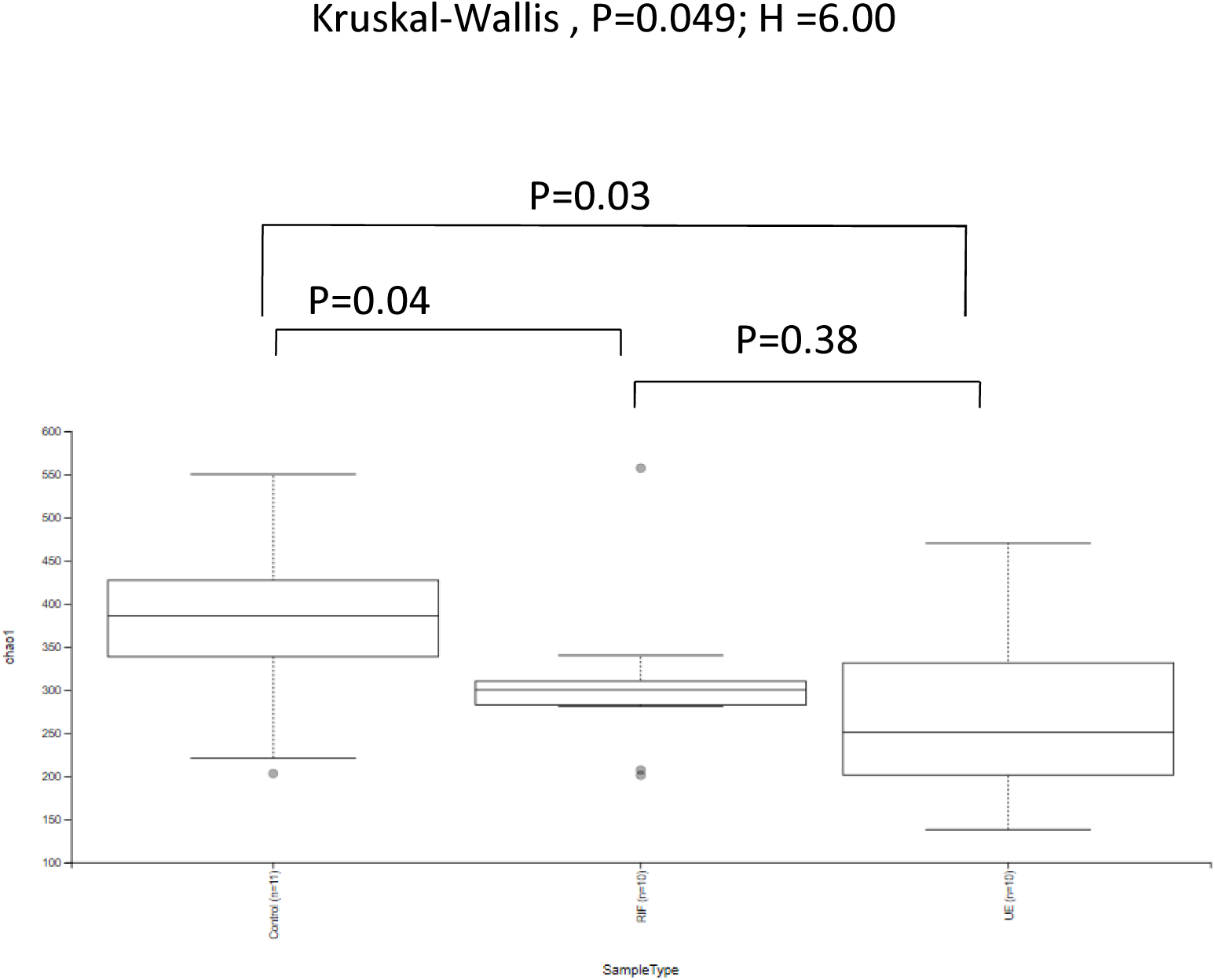
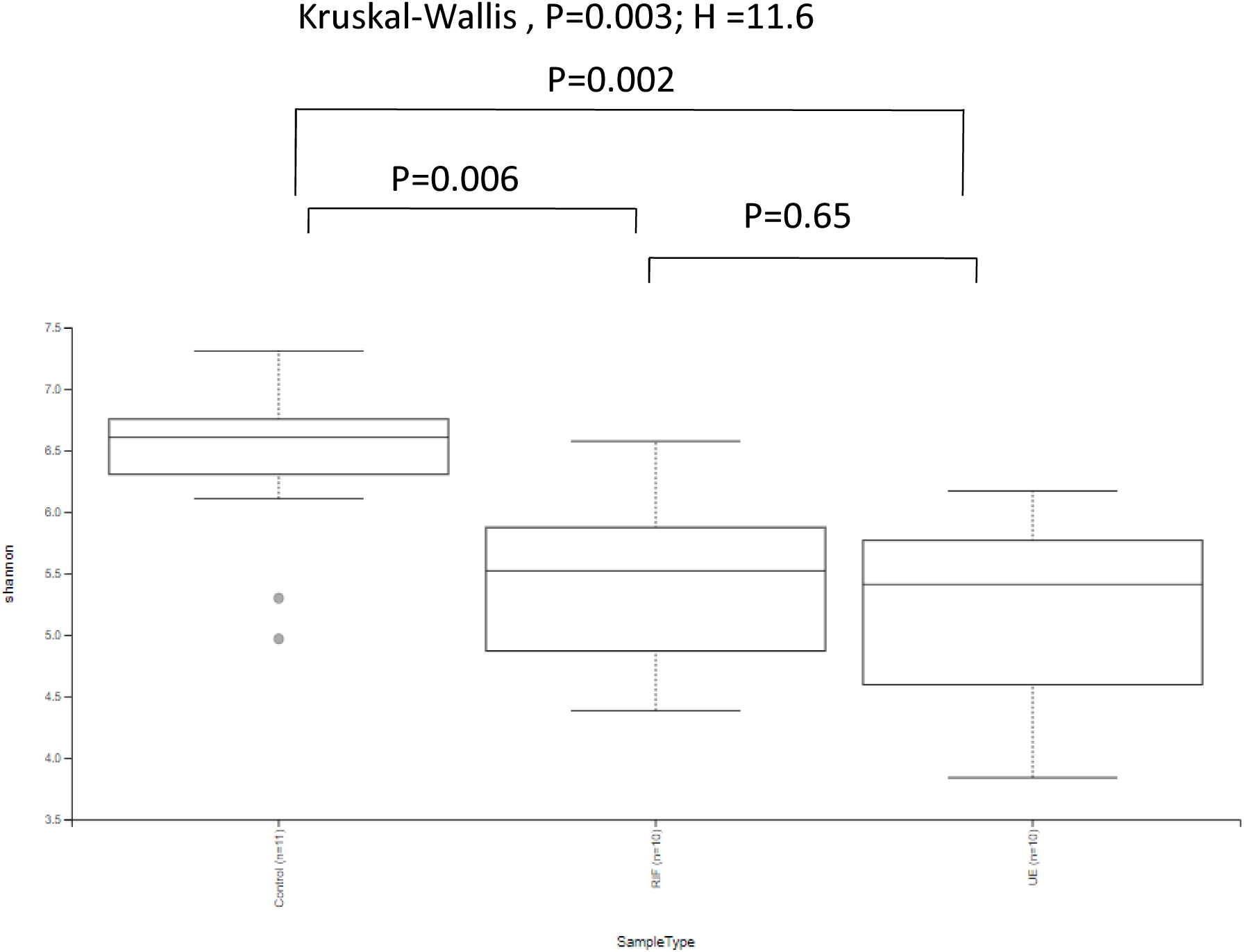
Box plots of α-diversity indices of the gut bacterial microbiomes of the control (CON, N = 11), RIF (RIF, N = 10), and the UE (UE, N=10) groups: (a) Shannon and (b) Chao 1 indices. The *P* values were determined by the Kruskal-Wallis test.

**Figure 2.**
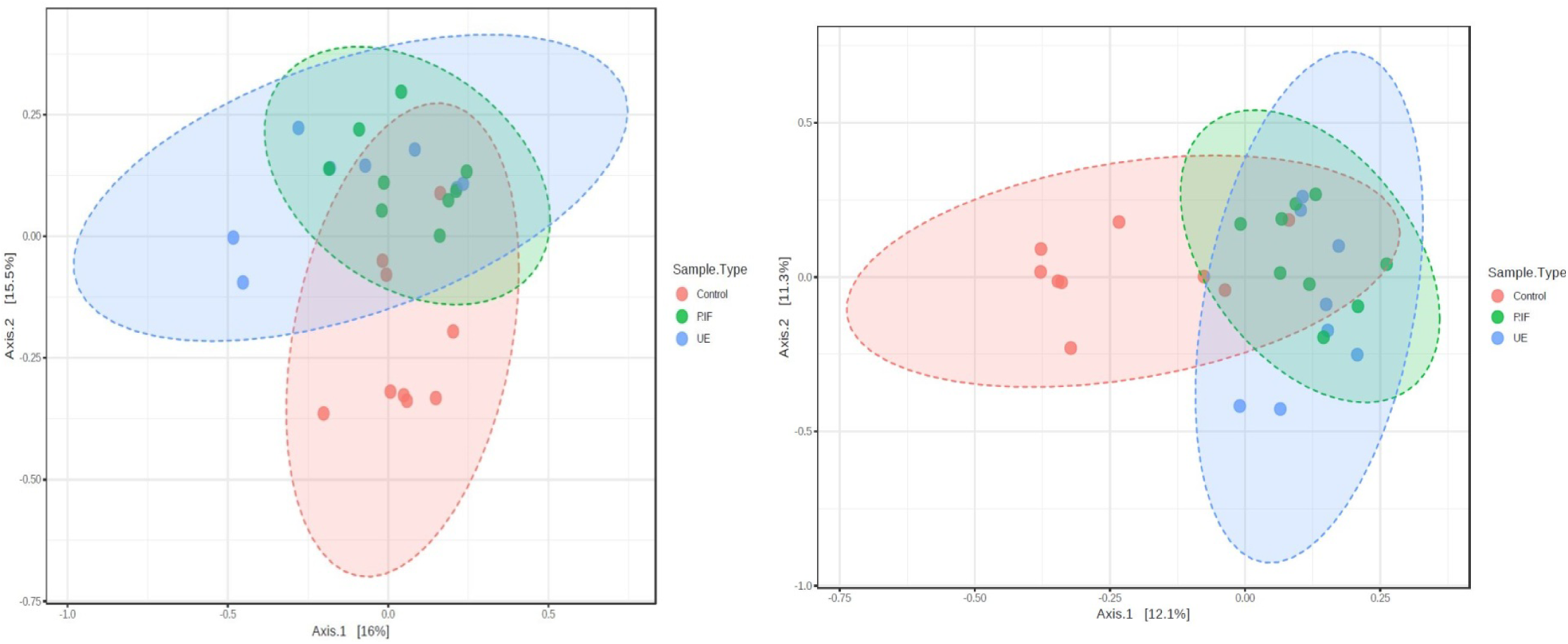
Differences in community composition (β-diversity) between the control (CON, Red, N = 11), RIF (RIF, Green, N = 10), and the UE group (UE, Cyan N=10) groups. Comparisons are based on the PCoA plots of Bray-Curtis (left) and Jaccard distances (right). Each principal coordinate axis represents the proportion of variance. Non-parametric multivariate analysis of variance (PERMANOVA) was used to calculate statistical differences between the groups. In the Bray-Curtis-based PCoA, the first and second axes of the PCoA accounted for 21.5% of the total variance (PERMANOVA, P< 0.05, R2 =0.12). In the Jaccard based PCoA, the first and second axes explained 23.4% of the total variance (PERMANOVA, P<0.05, R2 =0.10).

**Figure 3.**
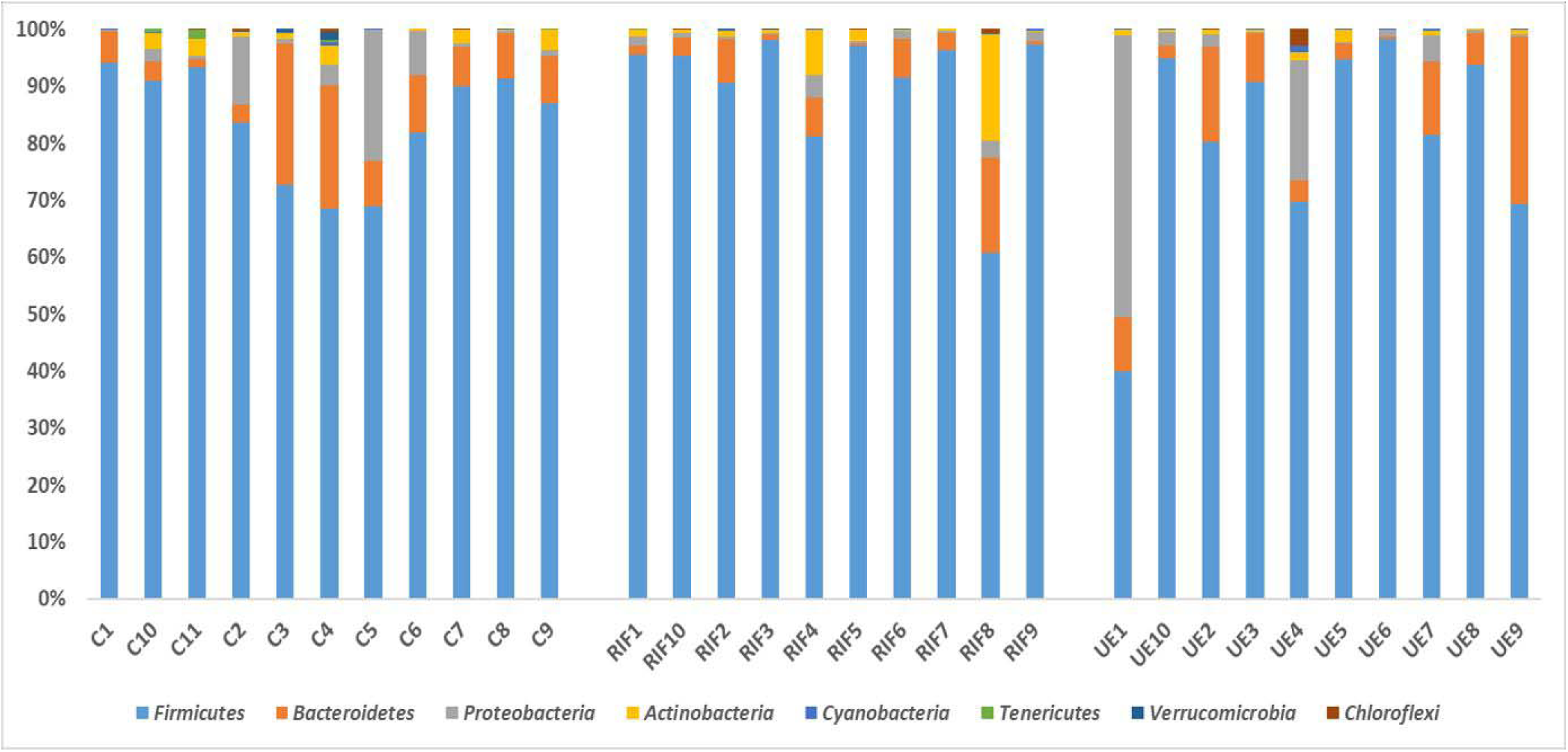
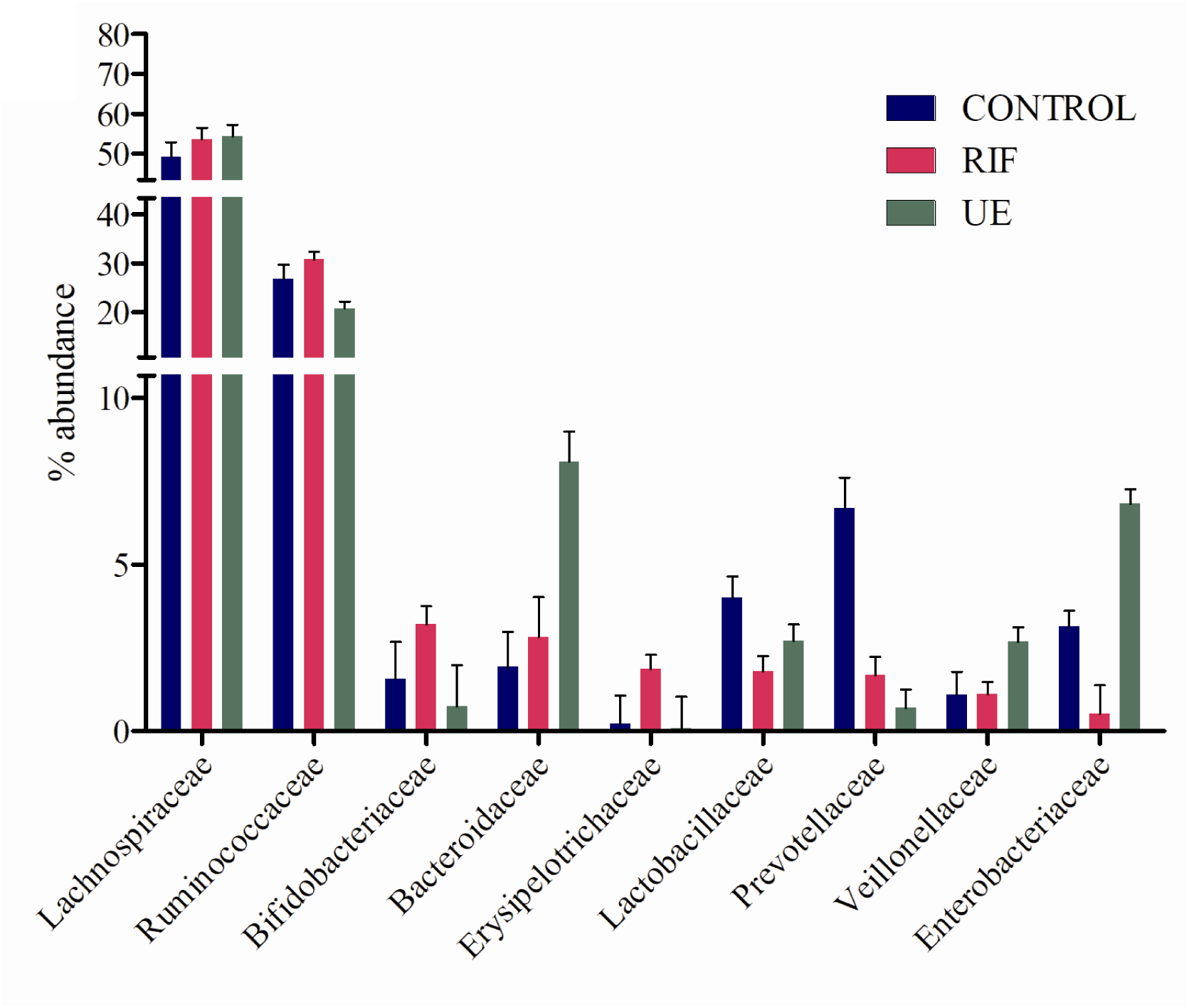
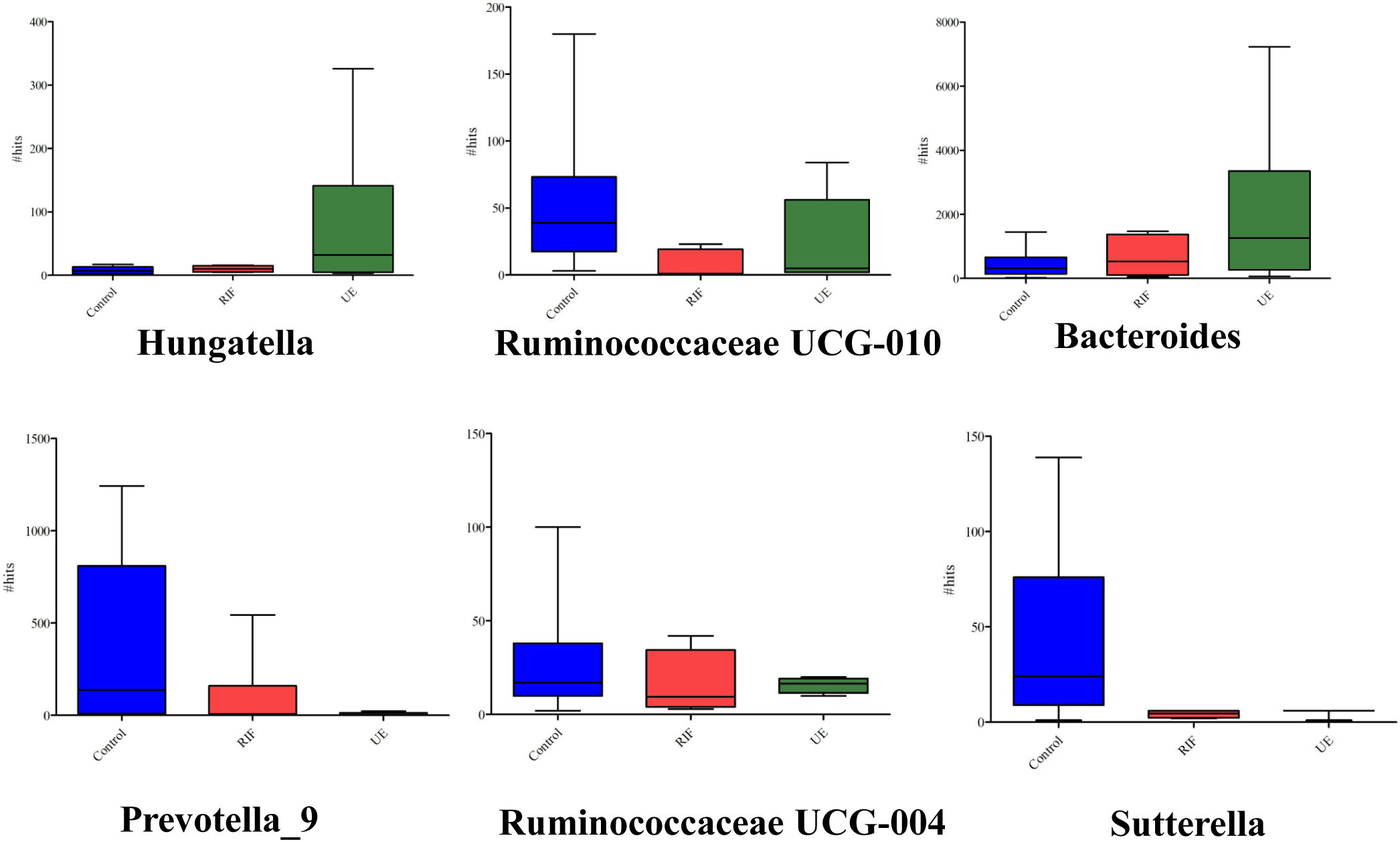
The bar chart shows the comparisons of relative abundances of top gut bacterial taxa between control (CON, N = 11), RIF (RIF, N = 10), and the UE (UE, N=10) groups at (a) the phylum (b) family and (c) genus levels. No differences in relative abundance were found between the phylum and family levels (P>0.05, Kruskal-Wallis test). Six bacterial genera showed statistically significantly (P<0.05, Kruskal-wallis test) differential abundance across the three groups are shown.

Regarding the bacterial community structure differences, the PCoA plot of the Bray-Curtis and Jaccard dissimilarity showed that bacteria of the RIF and UE groups overlapped and that both groups differed markedly from the controls **(Figures 2(a), (b))**. In the PCoA plot based on Bray-Curtis distances, the first and second axes of the PCoA explained 21.5% of the total variance with a significant difference (PERMANOVA, P< 0.05, R2 =0.12; **Figures 2(a)**). Showing the similar clustering pattern, in the Jaccard based PCoA plot, the first and second axes explained 23.4% of the total variance with a significant difference (PERMANOVA, P<0.05, R2 =0.10; **Figures 2(b)**).

### 3.5 Taxonomic analysis of Gut bacteria

After excluding the sequences that were present in less than 3%, we clustered the high-quality sequences into OTUs and assigned taxonomic identities. Consequently, we found 550 OTUs pertaining to 481 genera, 265 families, 156 orders, 68 classes, 715 species and 28 phyla. To evaluate the contribution of different taxa to diversity and composition, we calculated the relative abundance of taxa at the phylum, family and genus levels. Except at the genus level, we could not find statistically significant alteration of particular taxa at any other levels. Though, noticeable trends from a clinical point of view were still apparent in the data.

Across all 31 participants, the phyla level composition, represented by read percentages, showed usual human microbiota structures. Overall, among the detected 28 phyla, the four most abundant microbes were *Firmicutes* (85.10%), *Bacteroidetes* (7.70%), *Proteobacteria* (4.75%), and *Actinobacteria* (1.8%) **(Figure 3(a))**. With a relative abundance of less than 1%, the remaining bacterial population belonged to four phyla, including *Verrucomicrobia, Tenericutes, Cyanobacteria, and Chloroflexi*.

Firmicutes were abundant (7-9 %) in the RIF group than both the control and UE groups. *Firmicutes* were only slightly more abundant (2%) in the UE group than the controls. *Bacteroidetes* were depleted (50% less) in the RIF group than the other two groups. In contrast, the control and UE groups had the same levels of *Bacteroidetes. Proteobacteria* were relatively less abundant (4-7% less) in the RIF group than the control and UE groups **(Figure 3 (a))**. Among the three groups, the UE group had the highest levels of *Proteobacteria*, almost 2 fold higher than the controls. *Actinobacteria* were depleted in the UE group (∼4 fold less) compared to the other two groups, with the RIF group showing the highest abundance amongst all the groups, with 2 fold more abundance than the control group **(Figure 3(a))**.

The dominant bacterial families for all the subjects in the descending order of abundance were *Lachnospiraceae, Ruminococcaceae, Bifidobacteriaceae, Erysipelotrichaceae, Lactobacillaceae, Prevotellaacae, Vellinollacaea* and *Enterobacteriaceae* **(Figure 3(b))**. The levels of *Lachnospiraceae* and *Ruminococcaceae* were similar between the three groups. Notably, the levels of *Lactobacillaceae* and *Prevotellaacae* families were highest in the controls as compared to the other two infertile groups. *Bacteroidaceae, Vellinollacaea* and *Enterobacteriaceae* were highest in the UE group compared to the RIF and control groups, while *Bifidobacteriaceae* and *Erysipelotrichaceae* families were highest in the RIF group as compared to the control and UE groups **(Figure 3(b))**.

We further determined statistical differences in the specific bacterial genera of the three groups. Among 481 genera, the 6 were statistically significantly (p□<□0.05) different: *Bacteroides, Prevotella 9, Hungatella, Ruminococcaceae UCG-004, Ruminococcaceae UCG-010*, and *Sutterella*. Aside from *Bacteroides and Prevotella 9*, the abundance of the other 5 genera, while statistically significant (p□<□0.05), occurred in much lower proportions (<1%). Notably, *Bacteroides* and *Hungatella* were abundant in the infertile cohort, especially in the UE group, than the controls **(Figure 3(c))**.

*Bacteroides and Prevotella 9*, the two most highly abundant genera, varied significantly between the three groups. Compared to the controls, the relative abundance of *Prevotella 9* plunged noticeably in the RIF group (7 fold), and the UE group (8 fold), and the relative abundance of *Bacteroides* rose in the RIF group (1.5 fold) and UE group (3.2 fold) **(Figure 3(c))**. When we compared the infertile group against the controls, we found that in the infertile cohort *Prevotella 9, Ruminococcaceae UCG-004, Ruminococcaceae UCG-010* (p□<□0.05) declined, whereas *Bacteroides, Dorea, oral clone FR58* and *Peptoniphilus* increased (p□<□0.05) **(Figure 4S)**.

**Figure 4.**
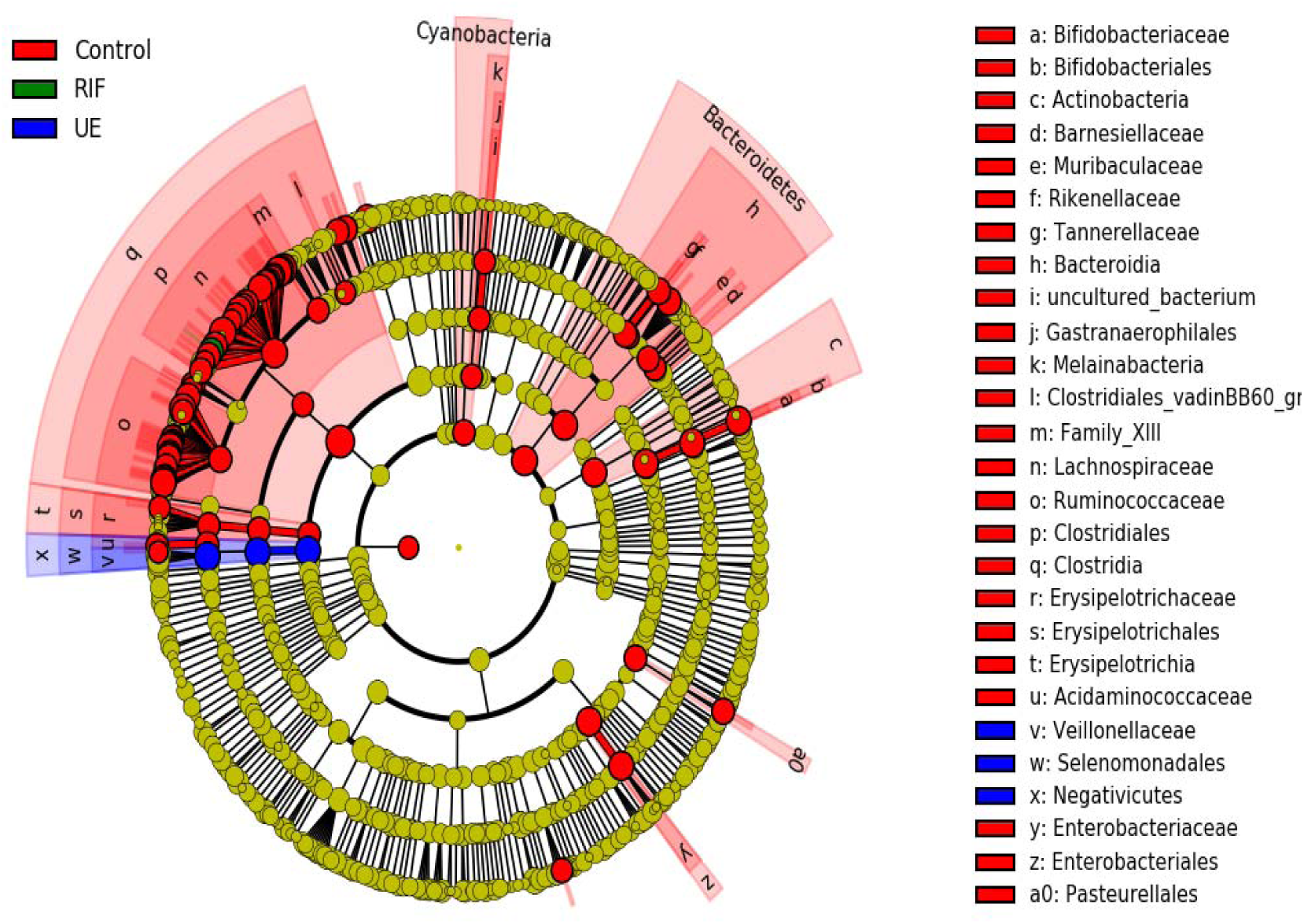
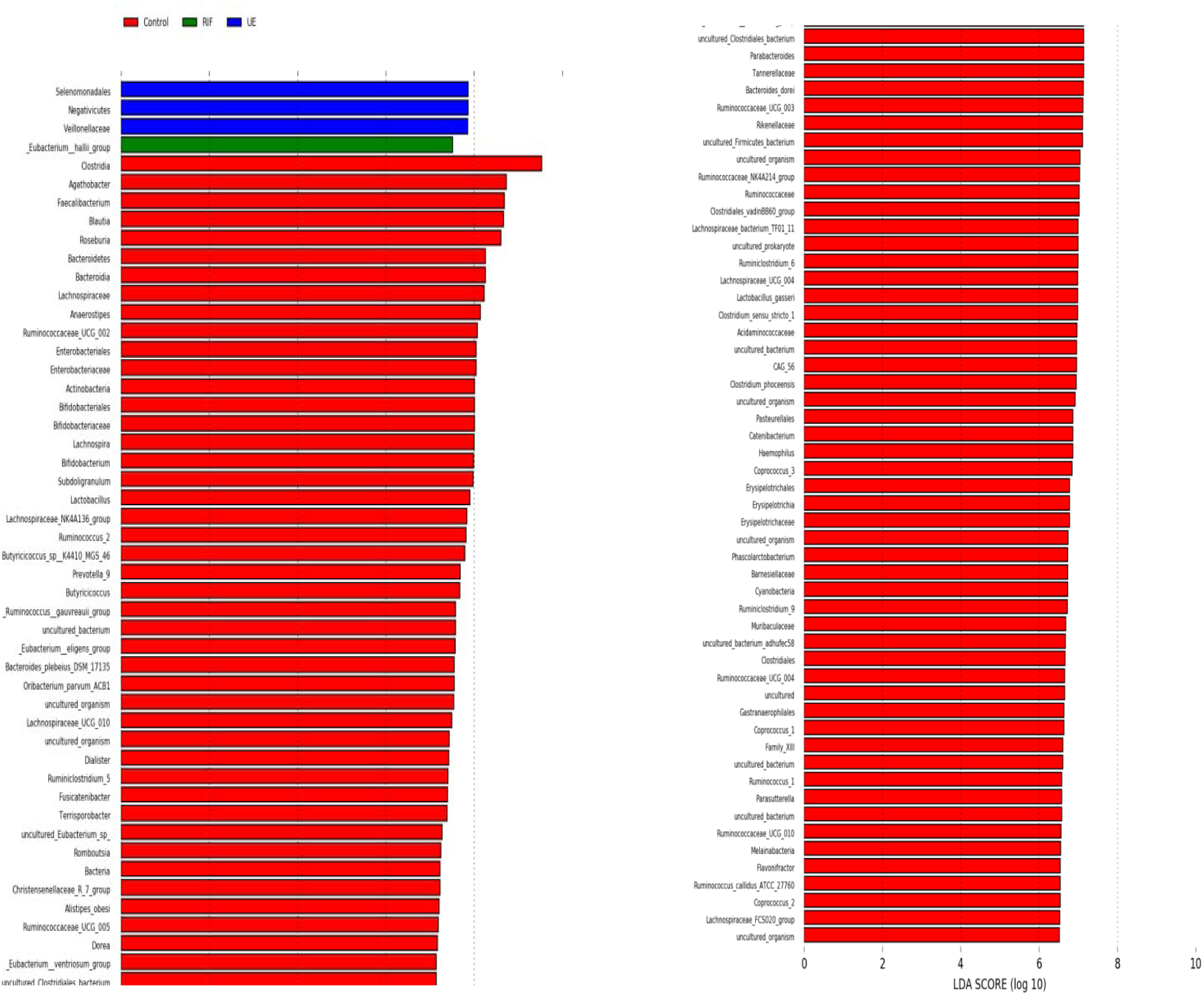
Distinct taxa of the gut bacteria determined by linear discriminant analysis effect size (LEfSe) analysis in the control (CON, N = 11), RIF (RIF, N = 10), and the UE (UE, N=10) groups. (a) The cladogram shows the taxa that were significantly elevated between the groups (b) Taxa with an LDA score significant threshold >3 are shown (P < .05; LDA score 3).

### 3.7 LEfSe analysis

LEfSe analysis was performed to assess the differentially abundant communities in the three groups **(Figure 4 (a), (b))**. In the controls, we observed diverse microbial communities with a high LDA score (Log10), with *Clostridia* showing the highest LDA score> 9 (p<0.05). In the RIF group, only *Eubacterium halli* showed the highest dominance with an LDA score>7 (p<0.05). In the UE group, unconventional *Firmicutes* such as *Veillonellaceae, Selenomonadales*, of the class *Negativicutes*, showed the highest preponderance with an LDA score > 7 (p<0.05).

### 3.8 Metagenomics of vaginal bacteria

Across the 24 vaginal samples, we found 384 distinct species belonging to 301 different genera classified in 135 different families, distributed into 10 phyla. We compared taxa between the three groups. Given the small sample size, we could not detect a significant statistical difference between them. Nonetheless, interesting trends in the data were apparent from a clinical point of view.

### 3.9 Taxonomic Analysis of Vaginal Bacteria

In the descending order, the dominant phyla, among the 10 detected phyla, included *Firmicutes, Fusobacteria, Proteobacteria, Actinobacteria, Bacteroidetes*, and *Patescibacteria* **(Figure 5(a))**. Of these, *Firmicutes* accounted for the vast majority of the vaginal bacteria in all the groups, with both the RIF (69%) and UE (69.71%) groups showing similar relative abundance, which was higher than the controls (53%). *Fusobacteria* (18% Vs. 0.07 Vs. 0.14) and *Bacteriodetes* (4.1% Vs0.17 Vs. 0.92) were relatively abundant in the controls than the RIF and UE groups. *Proteobacteria* were marginally more abundant in both RIF (15%Vs.11%) and UE (19% Vs. 11%) groups compared to the controls **(Figure 5(a))**.

**Figure 5.**
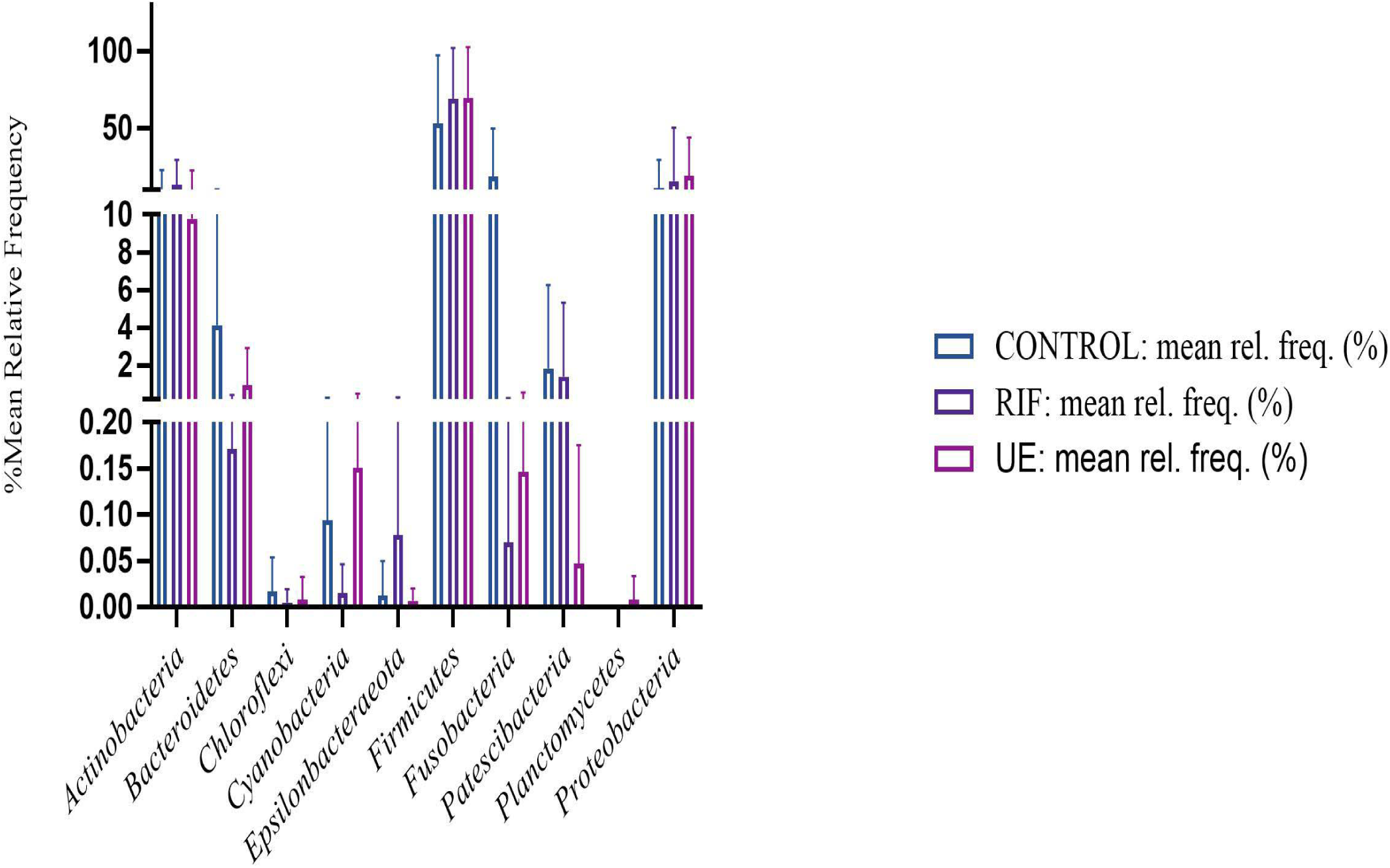
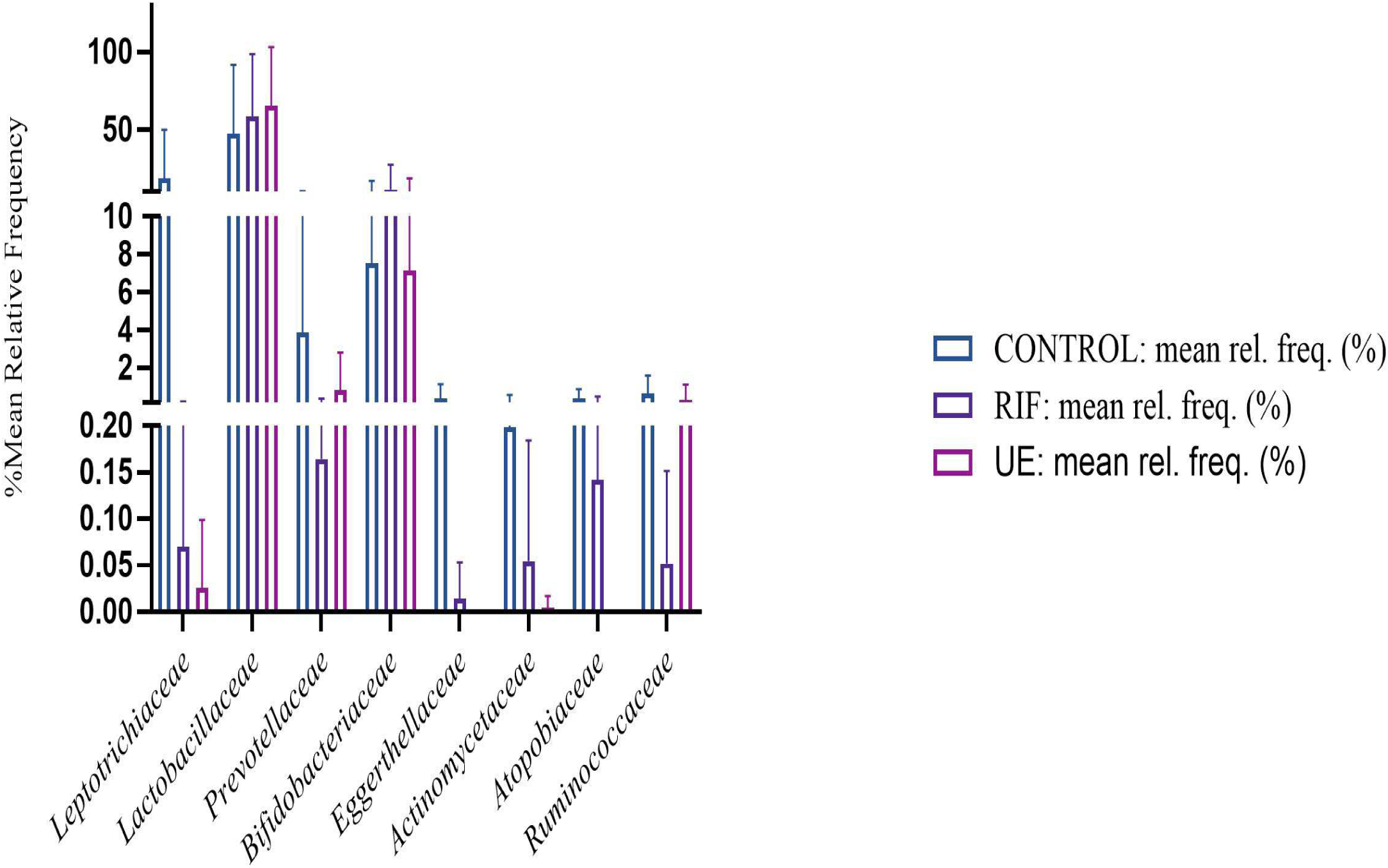
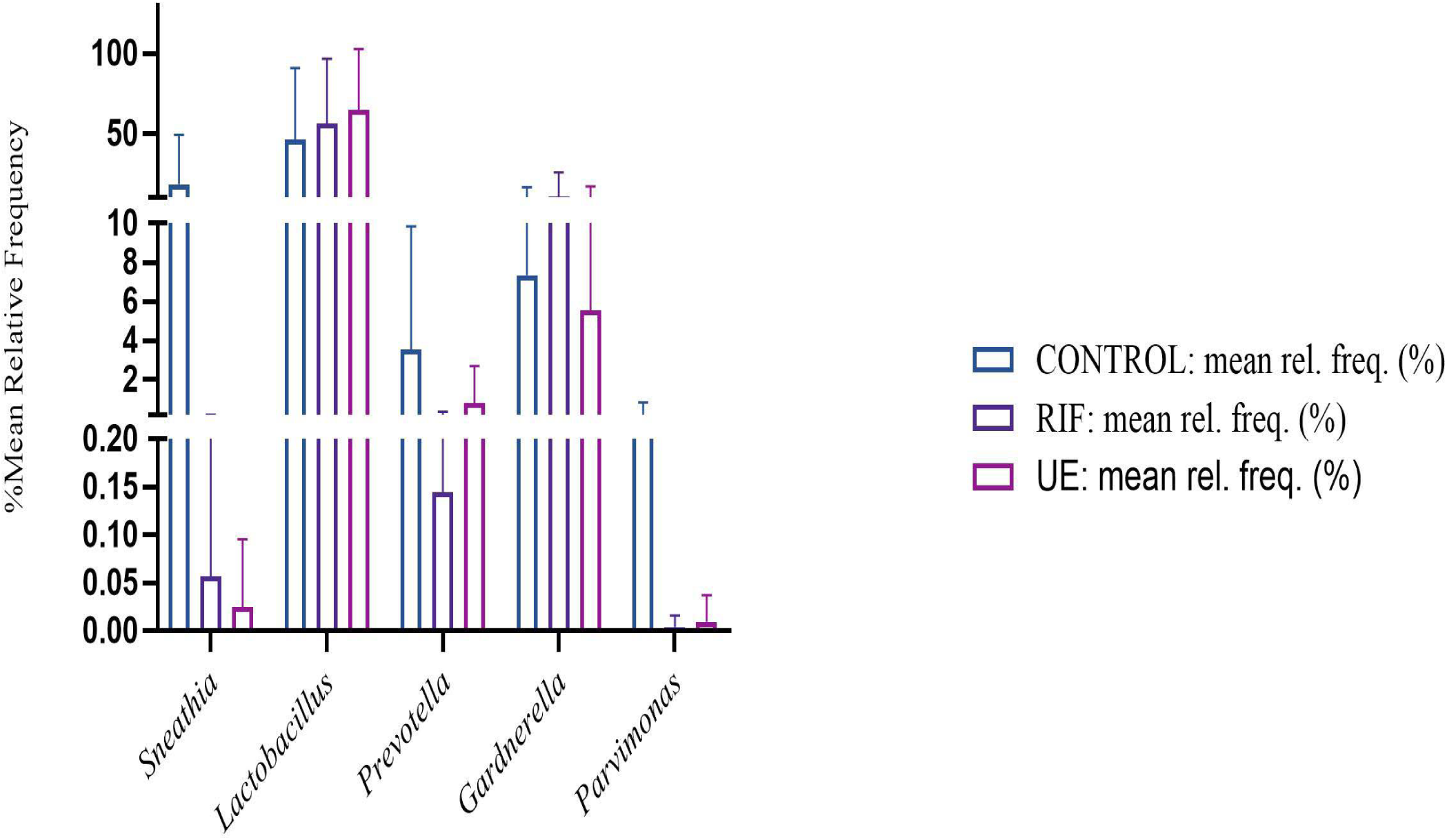
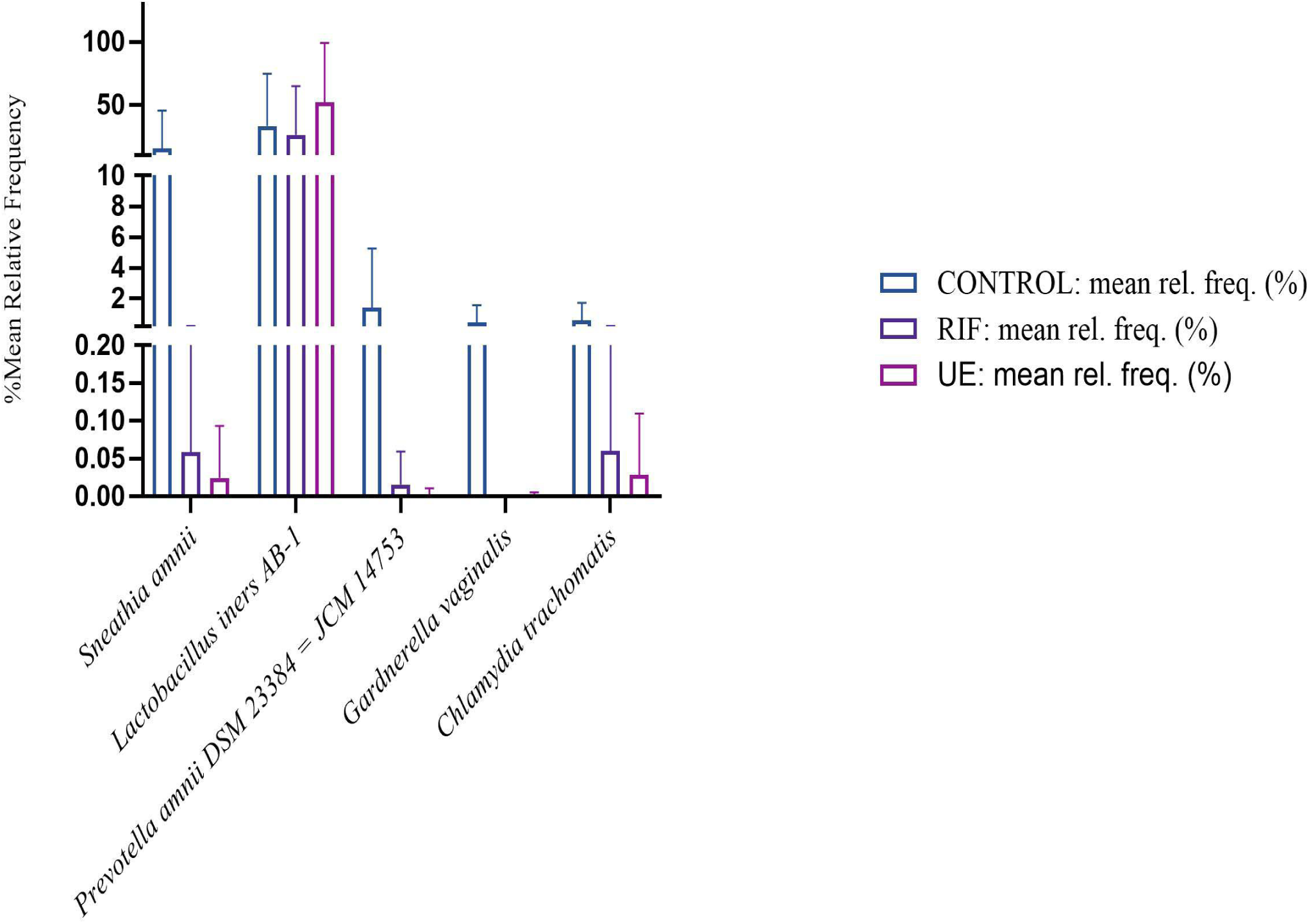
The bar charts show taxonomic comparisons of the vaginal bacteria between the control (CON, N = 8), RIF (RIF, N = 8), and the UE (UE, N=8) groups at (a) the phylum, (b) family (c) genus level (d) species levels. No differences in relative abundance were found at any levels (*P* > 0.05, Kruskal-Wallis test).

The dominant families for all the groups in the descending order of abundance were *Lactobacillaceae, Bifidobacteriaceae, Leptotrichiaceae*, and *Prevotellaceae* **(Figure 5(b))**. *Lactobacillaceae* and *Bifidobacteriaceae* were present in all the groups. Reflecting this trend at the phylum level, levels of *Lactobacillaceae* were higher in all the groups, with UE (65.3%) and RIF (58.41%) women showing the highest levels compared with the controls (47.2%).

At the genus level, 5 genera were detected, of which three were present in all the groups **(Figure 5(c))**. *Lactobacillus* was the most dominant genus among them, followed by *Gardnerella* and *Parvimonas. Gardnerella, Prevotella, Parvimonas*, and *Snaethia* were relatively more abundant in the controls compared to the infertile groups. Compared to the controls, *Lactobacillus* was relatively more abundant in the RIF and the UE groups (2-fold).

At the species levels, *Sneathia ammni* (0.36%) was detected only in the control groups. In contrast, *Lactobacillus iners AB-1*werepresent in all the groups, with descending order of relatively high abundance in the following manner: the UE group (62%), the controls (16%), and the RIF group (11.02%) **(Figure 5(d))**.

### 3.10 LEfSe analysis

We performed LEfSe analysis to assess the differentially abundant vaginal bacterial communities in the three groups. We could only find significant differences in the controls, with *Leptotrichia*, of the phylum *Fusobacteria*, showing an LDA (Log10) score >3 (p<0.05) **(Figure 6)**.

**Figure 6.**
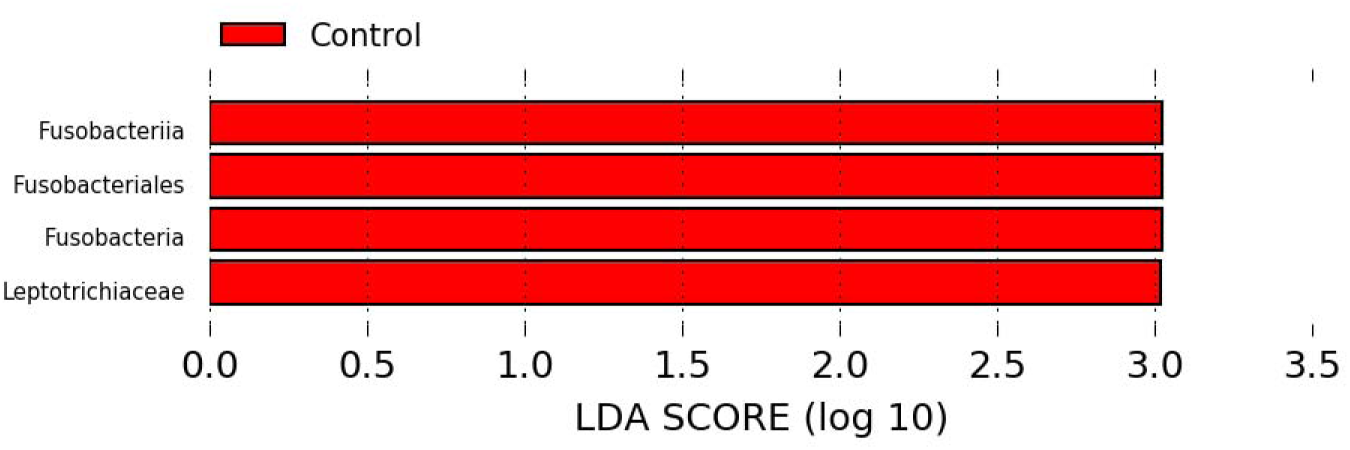
Differentially abundant vagina bacteria between the control (CON, N = 8), RIF (RIF, N = 8), and the UE (UE, N=8) groups as determined by linear discriminant analysis effect size (LEfSe) analysis. (a) Cladogram shows the taxa that were significantly enriched between the groups (b) Taxa with a LDA score significant threshold >3 are shown (P < .05; LDA score 3).

### 3.11 Alterations of the genus Lactobacillus Species

Within the genus of *Lactobacillus*, 9 species were identified. *L. gasseri, L. ruminis, and L. iners AB-1* were found in all the groups, with *Lactobacillus iners AB-1* being the most abundant species **(Figure 7)**. Among these, *L. jensenii* and *L. vaginalis* were only detected in the UE group, while *L*.*reuteri* was unique to the RIF group. *L. equicursoris, L. fermentum* and *L. salivarius* were unique to the controls.

**Figure 7.**
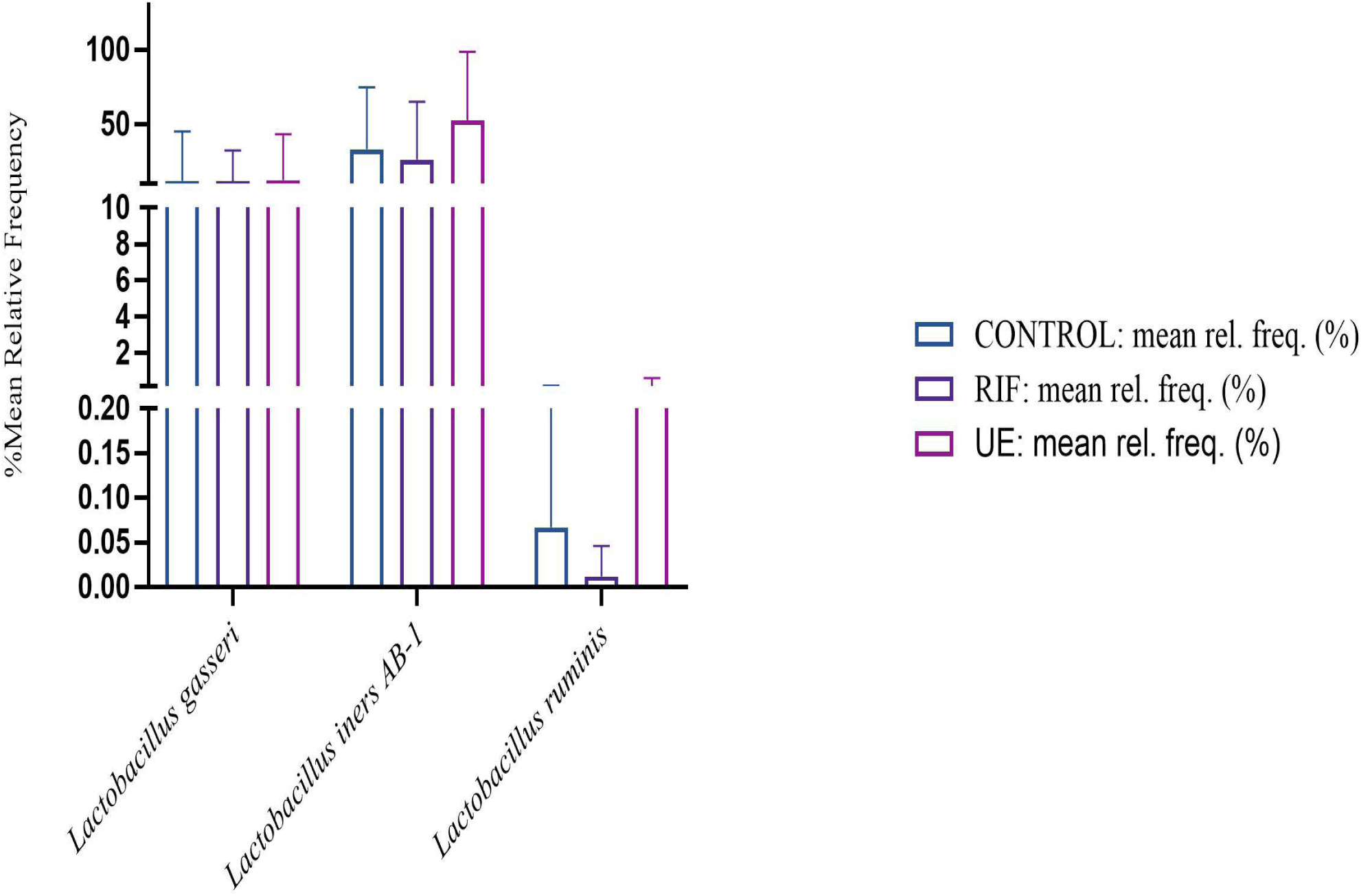
The bar charts show taxonomic comparisons of different *Lactobacillus* spp. of the vagina between the control (CON, N = 8), RIF (RIF, N = 8), and the UE (UE, N=8) groups. No significant differences in relative abundance were found between at any levels (*P* > 0.05, Kruskal-Wallis test).

## 4. Discussion

We compared, for the first time, the gut-vaginal microbiota axis of fertile women with that of women diagnosed with RIF and UE. The core findings include i) the infertile groups had gut dysbiosis as evident by low α-diversity indices and beta diversity metrics; ii) the gut microbial composition of the RIF and UE groups differed noticeably, with a set of Gram-positive taxa, mainly members of the phylum *Firmicutes*, being dominant in the former group and a set of Gram-negative bacteria, comprising members of the phylum *Proteobacteria* and the class *Negativicutes*, of the phylum *Firmicutes*, being dominant in the latter group; iii) butyrate-producing genera such as *Prevotella* declined in the infertile cohort; iv) elevated levels of the genus *Hungatella* occurred in the infertile cohort, especially in the UE; and v) the infertile cohort, especially the RIF group, had a comparatively healthy vaginal microbiota than the controls.

Gut microbial richness and diversity, defined by α-diversity indices, declined amongst the infertile groups, with the highest decline in the UE group. Studies have suggested that reduced α-diversity, an important indicator of gut microbiome health, indicates low-grade inflammatory disorders such as inflammatory bowel disease, metabolic disorder and obesity (Ott and Schreiber 2006, Le Chatelier, Nielsen et al. 2013, Al-Assal, Martinez et al. 2018). Of note, diet (vegetarian vs. Non-vegetarian) had little influence on α-diversity. Although, a fine-grained analysis showed vegetarian women had a higher proportion of the phyla *Patescibacteria* and the genus *Bacteroides*, likely reflecting its role in the digestion of plant fiber (Matijašić, Obermajer et al. 2014, Jang, Choi et al. 2017). Gut dysbiosis was also reflected in beta diversity indices, suggesting a distinct bacterial composition between the infertile cohort and the controls.

Notably, the reduction in the richness of gut bacteria in the RIF group was seen for the phyla *Bacteroidetes* and *Proteobacteria* compared with the other groups. Mirroring this, we observed a lower percentage of *prevotellaceae* (phylum *Bacteroidetes*), *Veillonellaceae* (phylum *Proteobacteria*), *Enterobacteriaceae* (phylum *Proteobacteria*). This was partly reflected at the genus level by a several-fold decrease in the genus *Prevotella* (phylum *Bacteroidetes*) and a mild elevation in *Bacteroidaceae* (phylum *Bacteroidetes*). In contrast, in the UE group, the reduction in richness was seen mainly for the phyla *Actinobacteria* and marginally for *Bacteroidetes*. In keeping with this, reduced levels of the *Bifidobacteriaceae* family (Phylum *Actinobacteria*) were noted, and a marginal decline in the phyla *Bacteroidetes* was reflected by the depletion of the *prevotellaceae* family (phylum *Bacteroidetes*) and a modest rise in *Bacteroidaceae* (phylum *Bacteroidetes*).

Altogether, the data clearly indicate that the reduced microbial richness in the RIF and UE groups is characterised by contrasting types of abundance at the taxa level. Nonetheless, data showed a partial overlap at the genus level in that a relative decline in the abundance of *Prevotella* and an increase in the abundance of *Bacteroides* commonly occurred in both the groups. Since *Prevotella* builds the protective gut mucosal barrier from short-chain fatty acids (SCFAs) such as butyrate and *Bacteroides* impedes mucin synthesis by producing metabolites such as succinate, acetate, and propionate, these findings suggest thinned mucosal protection is the common abnormality in the infertile cohort. (Mejía-León and Barca 2015). Of note, although not highly abundant in the infertile cohort, *Ruminococcaceae UCG-004 and Ruminococcaceae UCG-010*, of the *Ruminococcaceae* family and the genus *Sutterella* declined as well. These are also butyrate-producing genera (Vital, Karch et al. 2017, Jennings, Koch et al. 2019, Wang, Wichienchot et al. 2019).

When the mucus barrier is thinned, the gut bacteria and other microbe-associated molecular patterns (MAMPs) come in direct contact with toll-like receptors (TLRs), located in the gut epithelial cells such as the Paneth cells (Uchida, Oyanagi et al. 2014, Okumura and Takeda 2017, Steimle, Michaelis et al. 2019). TLRs recognise microbes and MAMPs, and subsequently elicit an immune response, leading to localised and systemic inflammation, which can cause implantation failure (Uchida, Oyanagi et al. 2014, Mejía-León and Barca 2015, Okumura and Takeda 2017, Steimle, Michaelis et al. 2019).

We proffer the following hypotheses to explain how the gut dysbiosis in the infertile groups causes implantation failure by separate mechanisms that promote systemic inflammation. We posit that gut dysbiosis–induced metabolic dysregulation plays a role in RIF. At the phyla level, abundances of *Bacteroidetes* and *Proteobacteria* were lower. By contrast, *Firmicutes* and *Actinobacteria*’s levels were higher in the RIF group compared with the other two groups--indicating an obesity-associated microbiota profile (Koliada, Syzenko et al. 2017). Obesity has been linked to an increase in *Firmicutes* to *Bacteroidetes* (F/B) ratio (Verdam, Fuentes et al. 2013, Koliada, Syzenko et al. 2017).

Conversely, weight loss has been shown to normalise this ratio (Koliada, Syzenko et al. 2017). The F/B ratio was elevated in six out of ten women in the RIF group. In contrast, only two to three subjects had elevated F/B ratio in the other groups. Unsurprisingly, the RIF group’s mean BMI, highest among the three groups, was in the obesity range (obesity ≥25 kg/m^2^ for Asian Indians (Mahajan and Batra 2018)), which chimes with the fact that obesity is a risk factor for RIF(Zhang, Liu et al. 2019).

Notably, the Clostridium XIVa cluster, of the *Firmicutes* phylum, whose members comprise flagellated bacteria with a tendency to colonise mucus, play a critical role in metabolic dysregulation such as obesity (including visceral adiposity) (Lopetuso, Scaldaferri et al. 2013, Verdam, Fuentes et al. 2013, Jennings, Koch et al. 2019). Indeed, a trend towards an increase in the relative abundance of *Firmicutes* genera in this cluster such as *Lachnoclostridium, Dorea, Ruminococcus 2, and Eubacterium* was duly noted in the RIF group (Lopetuso, Scaldaferri et al. 2013). Crucially, LEfSe analysis found that *Eubacteriumhalli*, a member of this cluster, previously found to be elevated in human obesity, is a RIF biomarker (Sanz, Santacruz et al. 2008, Verdam, Fuentes et al. 2013). Strikingly, the RIF group had the highest levels of the *Erysipelotrichaceae* (phylum *Firmicutes*) family. In contrast, it was almost absent in the two groups. Elevated levels of *Erysipelotrichaceae* has been linked to human obesity and has been correlated to elevated levels of Tumor Necrosis Factor-alpha (TNF-α), a pro-inflammatory cytokine involved in obesity-linked insulin resistance (Kaakoush 2015). Tellingly, a high relative abundance of *Firmicutes* has been shown to correlate with increased levels of peripheral TNF-α (Orbe-Orihuela, Lagunas-MartÃ-nez et al. 2018). A rodent study found that a high-fat diet first increased the phylum *Firmicutes*, corresponding with the changes of Paneth cell-antimicrobial peptides, which was later followed by the elevations of circulating inflammatory cytokines, including TNF-α, thus establishing causality between the phylum *Firmicutes* and TNF-α (Guo, Li et al. 2017).

Therefore, precisely, we postulate that the phylum *Firmicutes* generates TNF-α-driven systemic inflammation, and consequent insulin resistance may cause RIF. Strikingly, investigators showed elevated TNF-α/IL-10 ratio correlates with an increased risk of IVF failure (Winger, Reed et al. 2011). Chan *et al*. found that insulin resistance reduces implantation rate in *in vitro* maturation-*in vitro* fertilization-embryo transfer cycle (Chang, Han et al. 2013). Metformin, known to reduce the F/B ratio, has been shown to increase pregnancy rate in IVF repeaters without polycystic ovary syndrome (Jinno, Watanabe et al. 2010, Wang, Saha et al. 2018). Investigator showed an arginine-rich diet, known to reduce obesity and increase insulin sensitivity, corrects the elevated F/B ratio and increases embryo survival (Dai, Wu et al. 2011).

The most striking phyla level changed in the UE group involved depletion of *Actinobacteria* and abundance of *Proteobacteria*, the pro-inflammatory phylum, comprising common pathogens (e.g., *Escherichia, Salmonella*) (Sekirov, Russell et al. 2010). Compared to the controls, only marginal changes occurred in the levels of phyla *Firmicutes* and *Bacteroidetes* in this group, suggesting a critical role of *Proteobacteria* phyla in UE. This concurs with the fact that *Bifidobacterium*, a genus of the depleted phyla *Actinobacteria*, inhibits gut pathogens (Azad, Sarker et al. 2018). Unsurprisingly, compared to the other two groups, the UE group had the highest enrichment of pathogenic Gram-negative families, whose outer membrane contains a unique component, lipopolysaccharide (LPS) (Brown 2019). These bacterial families included: *Bacteroidaceae* (phylum *Bacteroidetes*), *Veillonellaceae* (phylum *Firmicutes*) and *Enterobacteriaceae* (phylum *Bacteroidetes*). Cogently, LEfSe analysis revealed members of the *Negativicutes* class—such as *Veillonellaceae, Selenomonadales*, which are atypical gram-negative *Firmicutes*, which possess lipopolysaccharides in the outer membranes— were biomarkers of UE (Vesth, Ozen et al. 2013).

The abundance of *Negativicutes* has been linked with an increase in the systemic levels of IL-6, the pro-inflammatory cytokine (Leite, Rodrigues et al. 2017). Along this line, enrichment of other Gram-negative species has been shown to increase plasma IL-6 levels (de Man, van Kooten et al. 1989, Leite, Rodrigues et al. 2017, Higuchi, Rodrigues et al. 2018). LPS of Gram-negative species, a pro-inflammatory endotoxin, binds to TLR-4 in the gastrointestinal mucosa, triggering an inflammatory cascade that causes localised NF-κB activation, ensuing the systemic secretion of IL-6 (Steimle, Michaelis et al. 2019).

Taken together, we proffer that in the setting of the enfeebled mucosal barrier, the overload of Gram-negative bacteria activates the gut innate immune system, generating IL-6-driven systemic low-grade inflammation, ultimately leading to UE. Indeed, Demir *et al*. found higher serum IL-6 levels, but not TNF-alpha levels, in women with UE than fertile women (Demir, Guven et al. 2009). Since elevated IL-6 levels impair various aspects of reproductive physiology, including LH secretion, LH-induced ovulation, and FSH-stimulated E2 and progesterone release, the gut bacteria-induced higher IL-6 levels may thus cause UE through these mechanisms (Demir, Guven et al. 2009).

In the infertile cohort, fascinatingly, elevated levels of *Hungatella*, producers of trimethylamine N-oxide (TMAO), which enhances thrombotic potential through platelet hyperreactivity, were found than the controls (Zhu, Gregory et al. 2016, Genoni, Christophersen et al. 2019). This data raises the possibility that an overactive coagulation system is a common mechanism of implantation failure. Since levels of *Hungatella* were highest in the UE group, this indicates an important role of thrombosis in UE. Indeed, Azem *et al*. found inherited thrombophilia plays a role in repeated IVF failures, particularly in the subgroup with UE (Azem, Maslovich et al. 2001).

Regarding the landscape of the vaginal microbiota, data showed that the vaginal bacterial community was less diverse than in the gut. Rarefaction analysis found that the RIF group had the lowest microbial diversity of the three groups, suggesting a healthy vaginal microbiota in the RIF group (Lokken, Richardson et al. 2019). We posit that through TNF-α-driven systemic insulin resistance, gut dysbiosis in the RIF group causes hyperglycemia and consequently increases glycogen levels in the vaginal epithelium, which is required for the maintenance of healthy microbiota (Carrara, Bazotte et al. 2009, Amabebe and Anumba 2018). Alternatively, adipose tissue-driven rise in peripheral estrogen may increase the glycogen content of vaginal epithelial cells (Lokken, Richardson et al. 2019). This chimes with the finding that obesity protects against vaginal dysbiosis (Lokken, Richardson et al. 2019). By contrast, the highest microbial diversity in the control group suggests vaginal dysbiosis. LEfSe analysis found *Leptotrichia*, an opportunistic pathogen of the female urogenital tract of the phylum *Fusobacteria*, in this group (Thilesen, Nicolaidis et al. 2007). In line with this, other pathogenic genera such as *Gardnerella, Prevotella*, and *Snaethia* were relatively more abundant in the controls compared to the infertile groups (Thilesen, Nicolaidis et al. 2007, Amabebe and Anumba 2018).

Consistent with previous research, *Firmicutes*, mainly *Lactobacilli* spp., constituted the bulk of total bacteria of all the groups (Madhivanan, Raphael et al. 2014, Madhivanan, Alleyn et al. 2015). Of the nine detected *Lactobacilli* spp., four belonged to the *L. delbrueckii*, subsp. of the *L. acidophilus* group (*L. iners, L*.*gasseri, L. jensenii, L. equicursoris*), three belonged to the *L. reuteri* group (*L. fermentum, L. reuteri, L. vaginalis*), and two belonged to the *L. salivarius* group (*L*.*ruminis, L. salivarius*) (Salvetti, Torriani et al. 2012). Of the three groups, the *L. reuteri* group comprises mostly obligate heterofermentative lactobacilli, consistent with the finding that compared to the women in the USA, Indian women housed a higher proportion of obligate heterofermentative lactobacilli, reflecting a need for metabolic plasticity and an adaptation against high pathogen load (Salvetti, Torriani et al. 2012, Madhivanan, Raphael et al. 2014, Madhivanan, Alleyn et al. 2015). Three *Lactobacillus* species dominated the vaginal microbiota across the groups: *L. iners, L*.*gasseri*, and *L*.*ruminis*, with *L. iners* being the most abundant, suggesting the existence of community state type 3 (CST 3) of the five human vaginal microbial communities (HVMC) as classified by Ravel et al. (2011) (Ravel, Gajer et al. 2011). To put this in context, *L. crispatus, L. jensenii, L. gasseri*, and *L. iners* occur most frequently in the healthy vagina worldwide, although substantial geographic, socio-economic and ethnic variations exist (Madhivanan, Krupp et al. 2008, Madhivanan, Raphael et al. 2014).

By lowering the vaginal PH<4 through lactic acid, generating bacteriocins and hydrogen peroxide (H_2_O_2_), or acting as a competitive inhibitor, *Lactobacilli* spp. protect the vagina from opportunistic pathogens (Kalia, Singh et al. 2020, Wang, Fan et al. 2020). Notably, though this protection varies according to strains based on their ability to produce D–lactic acid, which keeps PH<4, and antibacterial compounds (Wang, Fan et al. 2020). Most studies have shown that *L. iners*, lacking the ability to produce D-lactic acid, is less protective than other strains such as *L. crispatus* and *L. salivarius* (Pino, Bartolo et al. 2019, Kalia, Singh et al. 2020, Wang, Fan et al. 2020). Indeed, the highest relative proportions of *Lactobacillus iners AB-1* occurred in the UE group, suggesting a weakened resistance to colonisation by pathogens (Wang, Fan et al. 2020).

In sum, we illuminated gut and vaginal bacterial communities’ landscape, both at broader and finer levels, in implantation failure-associated infertile women and fertile women and offer conjectures to explain the data. Most importantly, these findings suggest that gut dysbiosis does not necessarily lead to vaginal dysbiosis. We hope that this study has laid the foundation of research on the link between the gut microbiota, the gut-reproductive microbiota axis, and implantation failure, which can lead microbiota-based diagnostic tools and therapeutic strategies.

Our study has a few limitations. First, since it is an underpowered single-center study, multi-center longitudinal studies with a large sample size are needed. Second, although we suggested the mechanistic hypotheses, we did not measure alterations in the immune system, hormones, platelet parameters and bacterial metabolic products such as short-chain fatty acids. Third, owing to the limited resolution of the 16S rRNA-sequencing technique, we could not identify what specific bacterial species or strains were involved. Fourth, we have not controlled for the factors that influence the vaginal microbiota such as menses, vaginal douching, and contraception. Fifth, we included two women with RPL in the RIF group. Finally, the functional significance of many species such as *peptoniphilus* remains undetermined in our analysis as the literature is scant on these genera. Hence, future investigations should address these shortcomings.

## Data Availability

The 16S rRNA gene sequencing data for all the gut and vaginal microbiota samples analyzed in this study have not been deposited with the National Center for Biotechnology Information.

## Acknowledgements

The authors wish to acknowledge the technical support of Nitin Savaliya, of Gujarat Biotechnology Research Centre (GBRC), for fecal and vaginal samples sequencing run. We are also grateful to Dr Yuvraj Jadeja, Dr Nipa Shah, Dr Nikita Daswani, Hansa Khrishti and Harmi Thakkar for supporting the clinical work.

## Authors’ contributions

BP conceptualized the idea, contributed to the design, made a few suggestions on the statistical methods, interpreted the data, and wrote the manuscript except for the metagenomics part of the method section. NP contributed to the design, patient identification, sample collection, ethical approval, financially supported the clinical work, presented a part of the work, and critically discussed and corrected the manuscript’s clinical aspects. NP collected fecal and vaginal samples, performed DNA isolation, sequencing, assisted in the analysis, drafted a part of the material and method section, and prepared some graphs and tables. SP collected fecal and vaginal samples, performed DNA isolation, library preparation of samples, and contributed to the correction of materials and method section. NN analysed the metagenomics data and contributed to the writing of the metagenomics section of the manuscript. RP sequenced fecal samples and corrected material and method section. CJ, the director of GBRC, provided financial and technical support and guidance for metagenomics sequencing and critically discussed the data. NP and MP contributed to ethical permission, patient selection and clinical data acquisition and critically discussed the data. All the authors provided criticisms and read and approved the manuscript.

## Supplementary figures

**Figure 1S.**
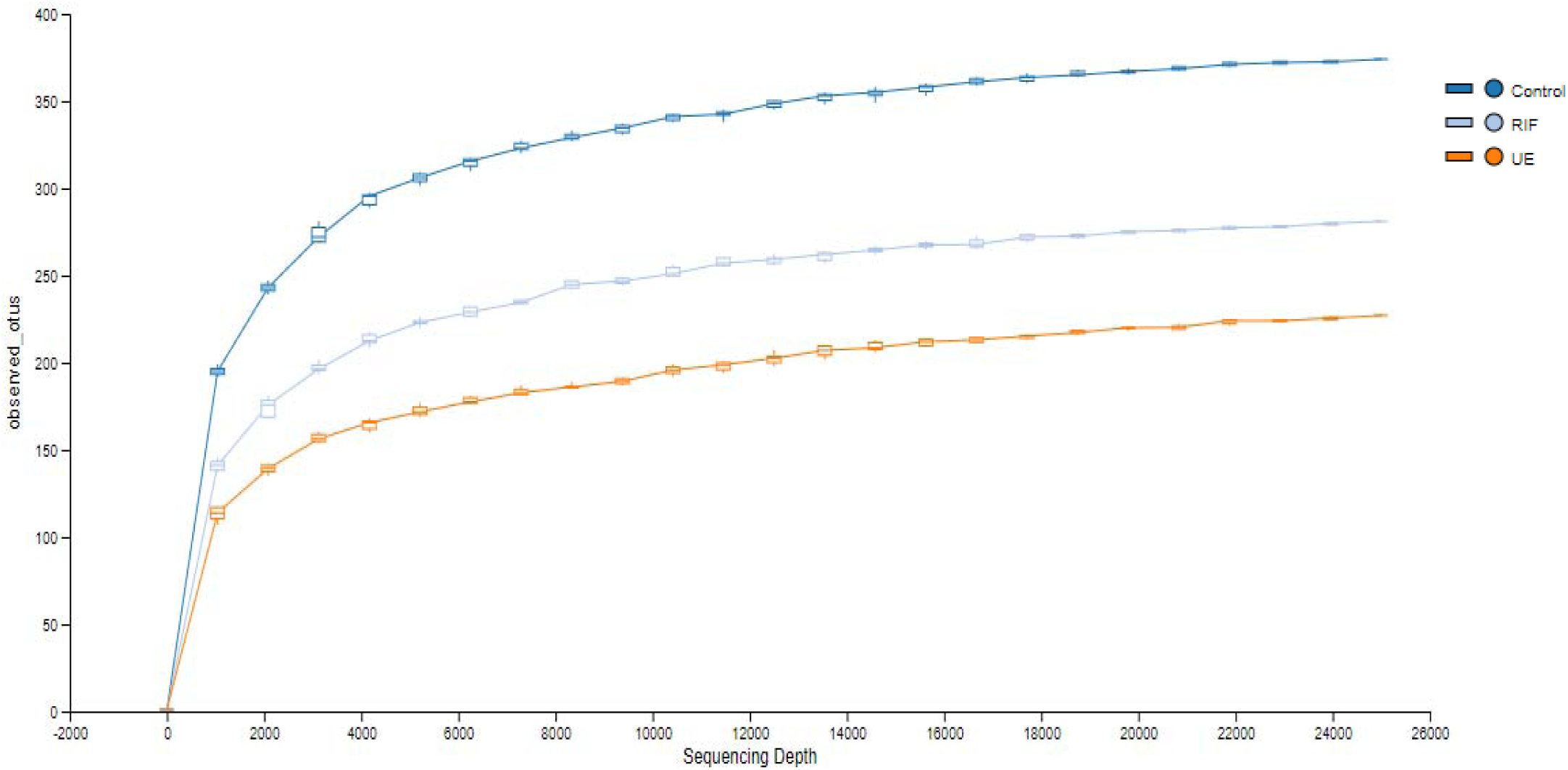
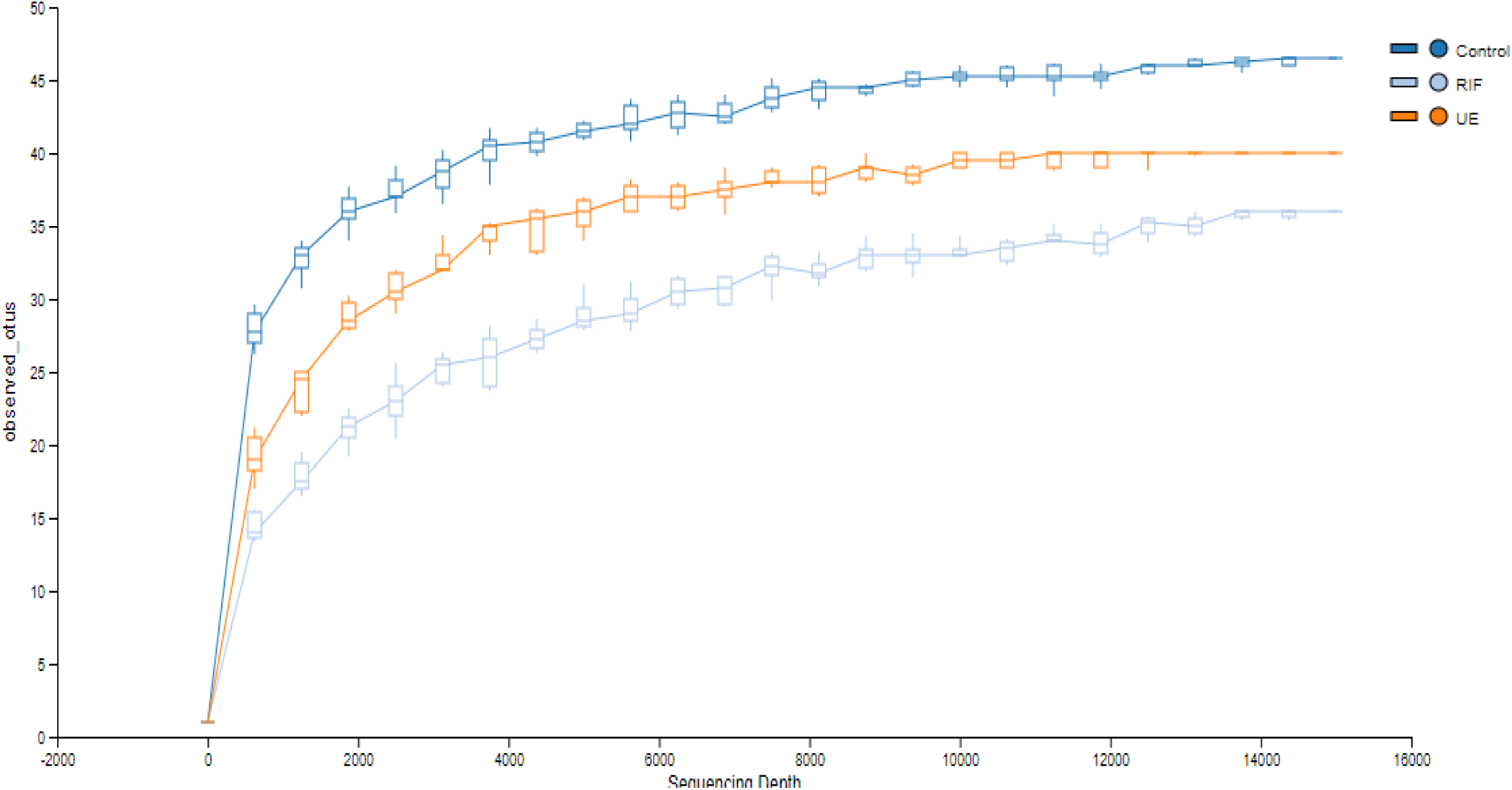
Rarefaction analysis of Gut and Vaginal Microbial diversity of the controls, RIF and UE groups. (A) Rarefaction curve of α-diversity of the gut bacteria in control (CON, N = 11), RIF (RIF, N = 10), and the UE (UE, N=10) groups (B) Rarefaction curve of α-diversity of the vaginal bacteria in control (CON, N = 8), RIF (RIF, N = 8), and the UE (UE, N=8) groups. The x-axis shows the number of sequences per sample, and the y-axis shows the rarefaction measure. When the curve plateaus, it shows that the sequencing data volume is sufficient to reveal most of the microbial information in the sample. The values of the y-axis reflect the community diversity of microbiota. The sequence number in the chart shows the sequence number of the sample. OTU, operational taxonomic unit.

**Figure 2S.**
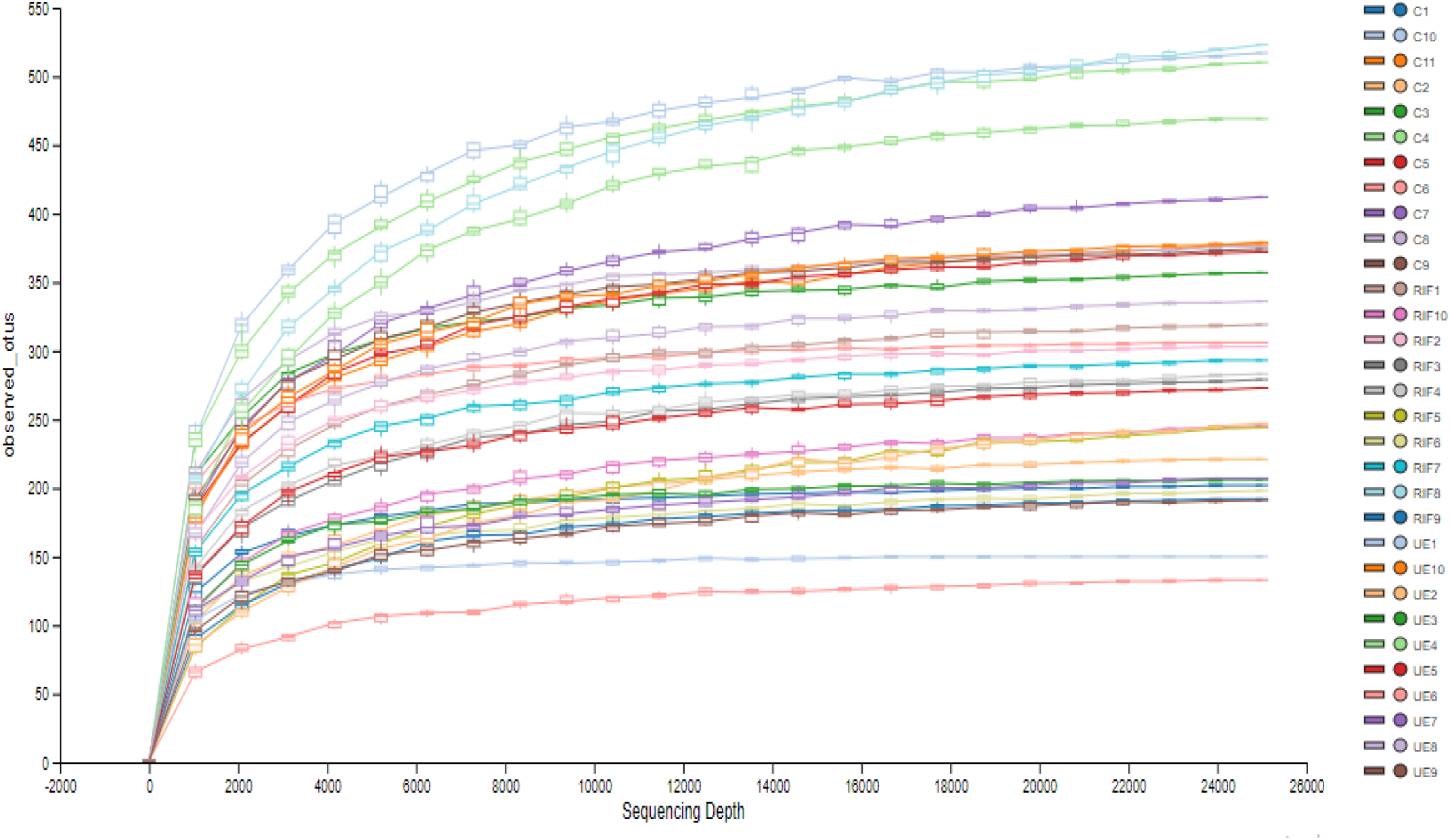
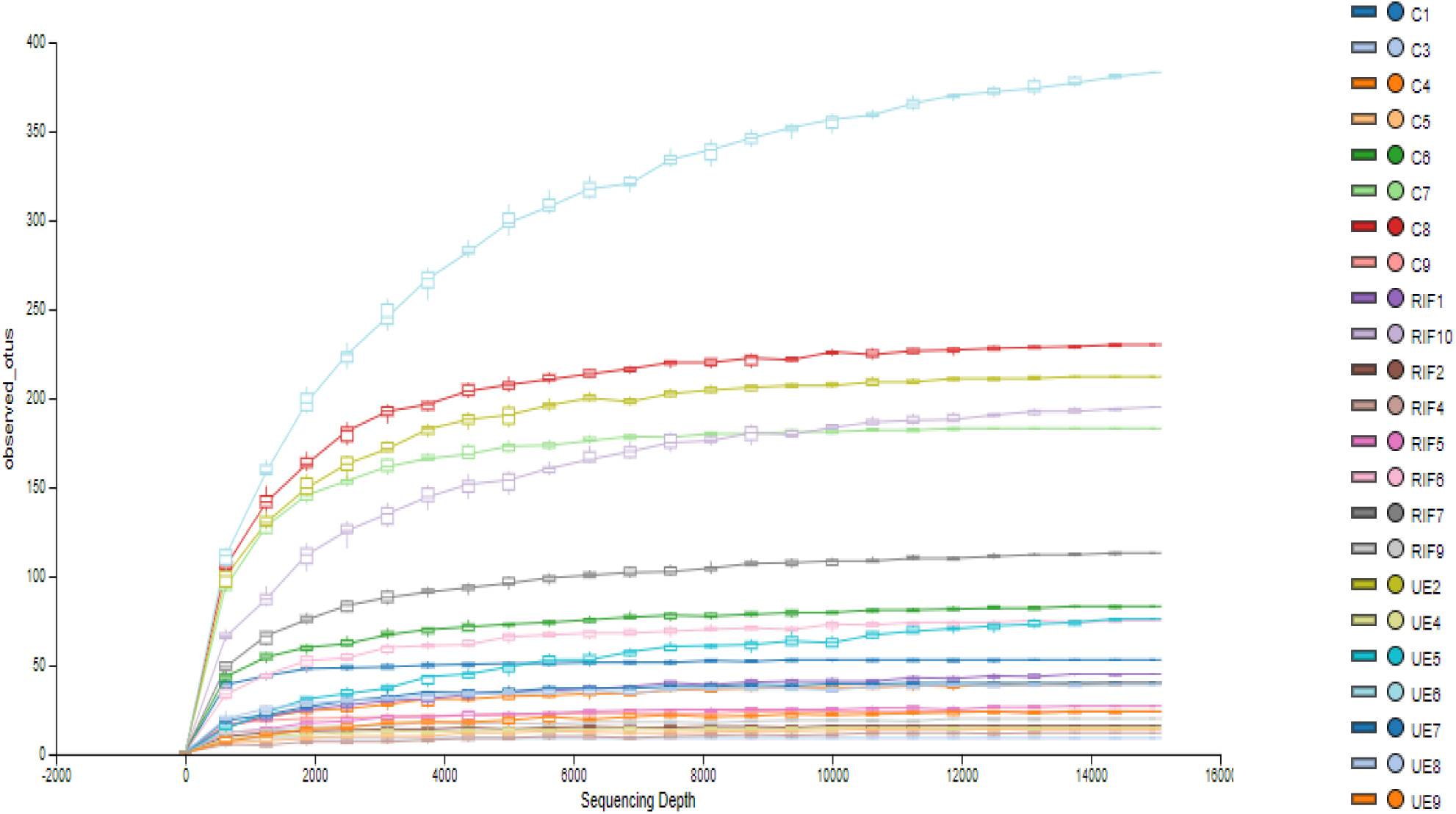
Rarefaction analysis of microbiome diversity sequences per sample: (a) gut (n=31) and (b) vaginal samples (n=24) (Operational taxonomic units (OTUs) for each sample at 97% of similarity). Each graph represents mean (column) and SD (bars).

**Figure 3S.**
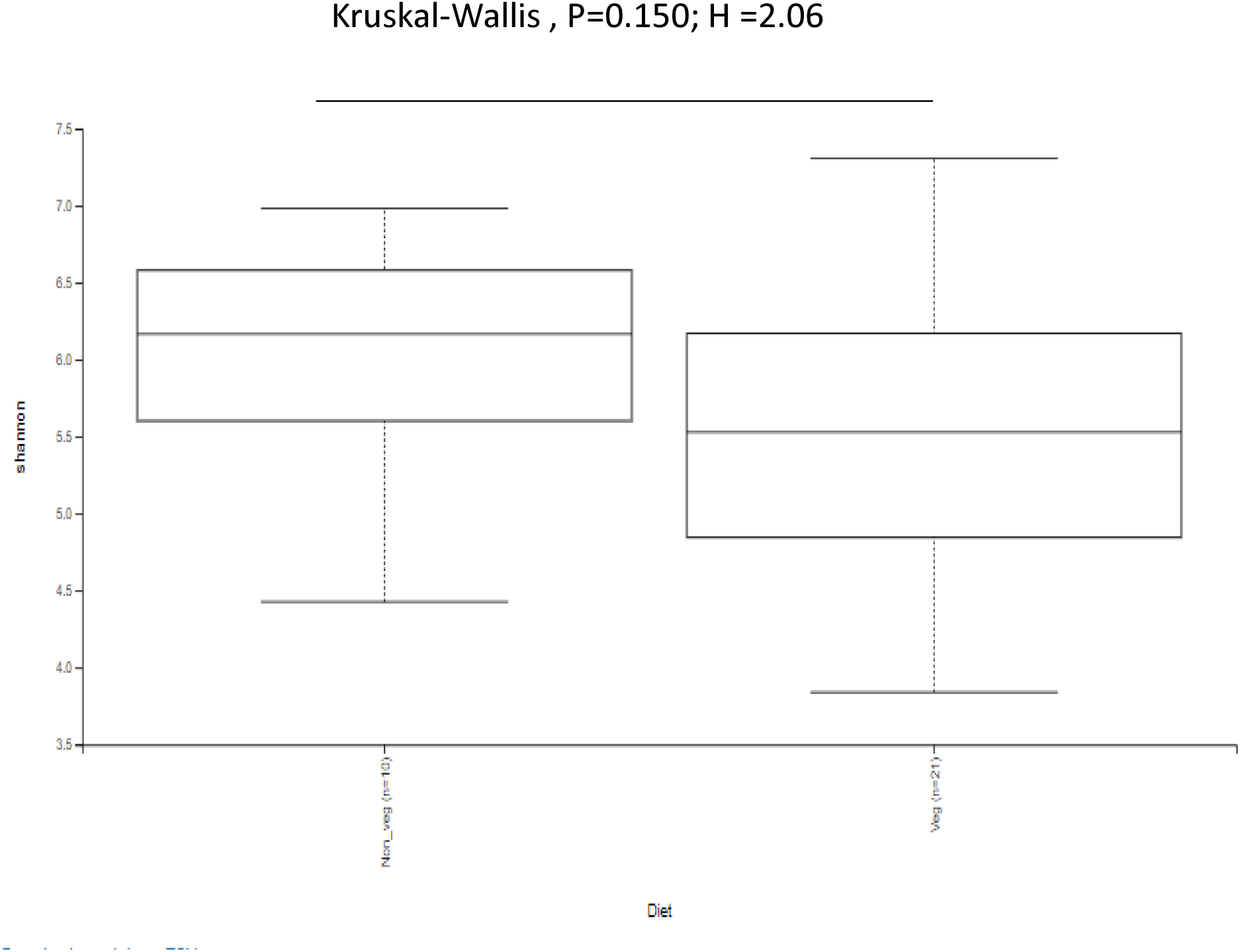
Influence of diet on α-diversity between vegetarian (n=21) and non– vegetarian participants (n=10). No significant difference in α-diversity between these groups was found (*P* > 0.05, the Mann-Whitney U test).

**Figure 4S.**
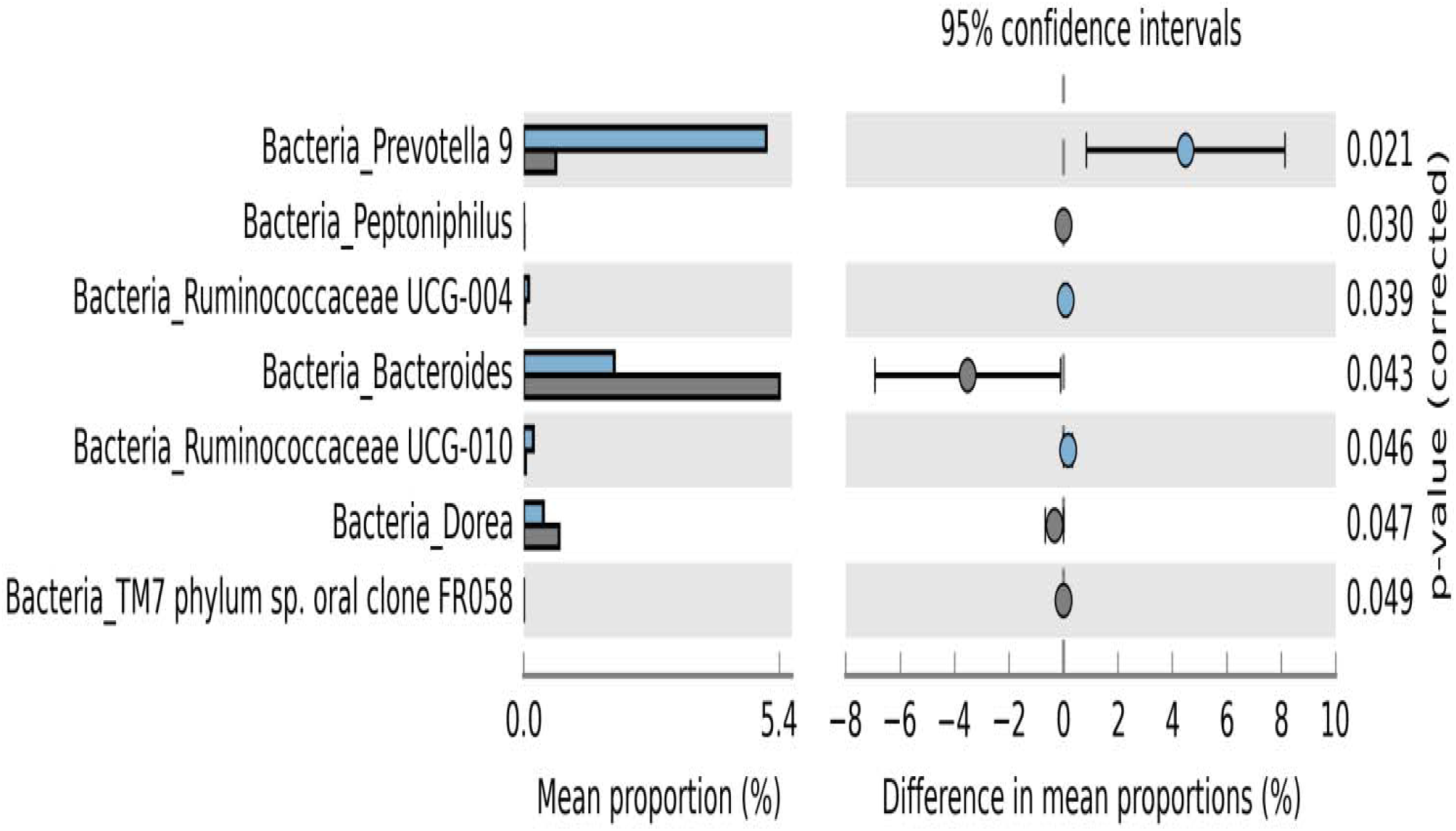
The bar chart shows taxonomic comparisons of the gut bacteria between the controls (CON, N=11) and the infertile cohort (the RIF plus UE groups, N=20) at the genus level with statistical significance values (*P* > 0.05, Mann-Whitney U test).

